# Impact of Participation Bias on Disease Prevalence Estimation in the *All of Us* Research Program: A Case Study of Ischemic Heart Disease and Stroke

**DOI:** 10.1101/2024.10.15.24315558

**Authors:** Younga Heather Lee, Ankita Patil, Cheryl R. Clark, Monik C. Botero, David W. Stein, Elizabeth W. Karlson

**Affiliations:** Psychiatric & Neurodevelopmental Genetics Unit, Center for Genomic Medicine, Massachusetts General Hospital, Boston, Massachusetts; Department of Psychiatry, Harvard Medical School, Boston, Massachusetts; Stanley Center for Psychiatric Research, Broad Institute of MIT and Harvard, Cambridge, MA; Department of Medicine, Brigham and Women’s Hospital, Harvard Medical School, Boston, Massachusetts; Department of Epidemiology, Harvard T.H. Chan School of Public Health, Boston, Massachusetts; Department of Obstetrics and Gynecology, Boston Medical Center, Boston University School of Medicine, Boston, Massachusetts

**Keywords:** Ischemic heart disease, stroke, ischemic stroke, coronary heart disease, heart disease, cardiovascular risk factors, social determinants of health, selection bias, epidemiology

## Abstract

**Importance:** Disease prevalence estimation is highly sensitive to sample characteristics shaped by recruitment and data collection strategies. Using follow-up study modules that require active participant engagement may introduce participation bias, affecting the accuracy of disease prevalence estimation.

**Objective:** To estimate the prevalence of ischemic heart disease (IHD) and stroke using electronic health records (EHR) and the self-reported Personal Medical History (PMH) survey collected in the *All of Us* Research Program.

**Design and settings:** Cross-sectional study aimed at estimating the prevalence of IHD and stroke among 266,472 participants with EHR in the latest release of the *All of Us* Registered Tier Curated Data Repository (R2022Q4R9).

**Main outcomes and measures:** Primary outcomes were IHD and stroke, ascertained using expert-curated diagnostic and procedure codes recorded in EHR. Secondary outcomes were IHD and stroke, ascertained using responses from the PMH survey. To mitigate the impact of participation bias in the PMH survey responses, we applied poststratification weighting based on annual household income and education.

**Results:** Of the 266,472 participants with EHR, 17,054 (6.4%) were identified as having IHD and 7,461 (2.8%) as having stroke based on the EHR definitions. Among PMH survey respondents, the EHR-based prevalence was lower at 5.6% (95% CI, 5.4-5.7) for IHD and 2.2% (95% CI, 2.1-2.3) for stroke, compared to 7.2% (95% CI, 7.0-7.3) for IHD and 3.3% (95% CI, 3.2-3.4) for stroke among non-respondents. The PMH survey-based prevalence among respondents was 5.9% (95% CI, 5.7-6.0) for IHD and 3.6% (95% CI, 3.5-3.7) for stroke, with higher estimates among non-Hispanic White participants after applying poststratification weights. **Conclusion and relevance:** Our findings suggest that while the current *All of Us* cohort with EHR reflects the general US population for IHD and stroke prevalence, participants completing the PMH survey are skewed toward higher socioeconomic status and medical literacy. Future research should refine bias mitigation strategies when using voluntary follow-up data to estimate disease prevalence in this cohort.

**Key Points:** *Question:* What is the prevalence of ischemic heart disease (IHD) and stroke in the *All of Us* Research Program cohort with electronic health records (EHR), and in the subset of these participants who also completed the Personal Medical History (PMH) survey?

*Findings:* The EHR-based prevalence estimates of IHD and stroke were 6.4% and 2.8%, respectively. They were significantly lower among PMH survey respondents but higher among non-respondents.

*Meaning:* Participants who complete follow-up study modules, such as the PMH survey, may disproportionately represent those with higher socioeconomic status and better health, potentially leading to an underestimation of IHD and stroke prevalence.

## Introduction

Ischemic heart disease (IHD) and stroke are the largest causes of mortality in the US, representing one in five deaths annually.^1^ The prevalence of these conditions varies by demographic factors, including race, ethnicity, and sex, with disproportionate deaths seen among people identifying as Black or African American, Hispanic or Latino, and among men.^2,3^ To capture such variations in the occurrence and course of illness across populations, it is essential to use data from cohorts reflecting the broad diversity of the US population.

The longitudinal *All of Us* Research Program has enrolled more than 750,000 participants from diverse enrollment sites across the United States, including regional medical centers, federally qualified healthcare centers, Veterans Administration hospitals, and through direct volunteer outreach.^4^ In addition to the unprecedented scale and diversity of the cohort, it provides a unique and valuable resource by collecting a wide range of health-related data modalities. They include, but not limited to, electronic health records (EHR), various participant-reported surveys (e.g. health history, lifestyle, pandemic experience), genomic, wearable—all gathered using a disease-agnostic approach to support a broad spectrum of health research.^5^ Aside from the Basics and Lifestyle surveys, other follow-up surveys are administered online several months after the initial enrollment and rely heavily on active engagement. Participation in follow-up survey varies by participant characteristics,^6^ such as age, gender, socioeconomic status, which can potentially lead to inaccurate representation of the experiences and health outcomes.

In the present study, we estimate the prevalence of ischemic heart disease (IHD) and stroke using EHR-based case definitions of among *All of Us* participants with EHR. We compared the EHR-based prevalence estimates between respondents and non-respondents of the PMH survey, hypothesizing that the EHR-based prevalence would be lowest among respondents and highest among non-respondents. Additionally, for respondents, we additionally calculated prevalence using survey-based case definitions. We then compared participant characteristics between PMH survey respondents and non-respondents to guide the specification of poststratification weights, aiming to mitigate the impact of participation bias. We hypothesized that the weighted PMH survey-based prevalence estimates for IHD and stroke would be higher than the unweighted estimates.

## Methods

### Study Population

At enrollment, participants are asked to consent to the release of EHR through the secure program database after the data transformation into a standardized Observational Medical Outcomes Partnership Common Data Model (OMOP CDM) format. As of March 2024, there were 266,472 participants (65.1% of the overall *All of Us* cohort) whose EHR were available for analysis (Registered Tier Dataset v7, version R2022Q4R9; released on April 20, 2023; see **eMethods 1** for details on recruitment and data collection). We then stratified our study sample based on the completion of the Personal Medical History (PMH) survey.

### Primary outcomes: EHR-based case definitions of IHD and stroke

We identified clinician-diagnosed IHD if participants had 2 or more qualifying ICD-9 or 10 diagnostic and/or procedure codes for IHD on separate days. We additionally identified cases of IHD using an expert-curated set of Current Procedural Terminology (CPT) codes for cardiac revascularization procedures. Separately, we identified clinician-diagnosed stroke cases if participants had 2 or more ICD-9 or 10 diagnostic and/or procedure codes for stroke on separate days. The complete list of qualifying codes for both IHD and stroke cases in the *All of Us* cohort, informed by and extending the previous study,^7^ can be found in **eTable 1**.

### Secondary outcomes: PMH-based case definitions of IHD and stroke

The “Personal Medical History (PMH)” and “Family History” surveys were collected three months after enrollment during the first two years of cohort follow-up. In 2019, these were combined into a single “Personal and Family History” survey and offered to participants at the baseline visit, with a reminder sent at three months to complete follow-up surveys. As secondary outcomes, we identified self-reported cases of IHD and stroke based on PMH survey responses from 124,192 participants (46.6% of the study sample). In this survey, participants are asked whether they were previously told that they have disease conditions from the predefined list of 11 comorbidity groups (shown in **eMethods 2**). In the present study, we focused the “Heart and Blood Conditions” section, in which participants were asked about coronary artery or heart disease, stroke, and transient ischemic attacks (TIAs). For IHD, participants who reported being previously diagnosed with coronary heart disease or heart attack were considered PMH-based cases. For stroke, participants who reported being previously diagnosed with stroke and TIAs were considered PMH-based cases.

### Covariates

We examined the 16 sociodemographic, lifestyle, and health literacy characteristics measured at enrollment for their potential associations with the likelihood of either releasing EHR or completing the PMH survey. Sociodemographic characteristics included age (in years), sex assigned at birth (female, male, neither male nor female), birthplace (USA, non-USA), self-reported race and ethnicity (Hispanic or Latino, Non-Hispanic (NH) Asian, NH Black or African American, NH White, NH more than one population, refused/skipped/none of these), region of residence (Northeast, Midwest, South, West), marital status (never married, married/living with a partner, divorced/separated/widowed), health insurance status (yes, no), employment status (employed for wages or self-employed, not currently employed for wages), highest level of education received (college or higher, less than college), household income (50k or higher, less than 50k), and current homeownership (own, rent, other arrangements). Lifestyle characteristics included lifetime cigarette smoking (yes, no) and lifetime alcohol consumption (yes, no). Lastly, we also included measures of medical literacy using the first three questions from the Overall Health Survey shown in **eMethods 3**.

### Statistical analysis

#### Prevalence estimation

We estimated EHR-based prevalence among 266,472 participants with linked EHR. Among 124,192 (46.6%) participants who also completed the PMH survey, we additionally estimated PMH survey-based prevalence. Lastly, we applied poststratification weighting to the PMH survey-based prevalence estimation, accounting for differences in household income and education between PMH survey respondents and non-respondents.

#### Poststratification weighting

We used two approaches to specify poststratification weights. We first performed univariate descriptive analysis to compare the distributions of 16 demographic, socioeconomic, and medical literacy characteristics between the overall study sample, PMH survey respondents, and non-respondents, using chi-square tests. We then conducted a multivariable logistic regression analysis to estimate the probability of completing the PMH survey among participants who released EHR. We triangulated the evidence from the two approaches to specify the most parsimonious set of covariates to calculate poststratification weights. After specifying the covariates, we used the survey R package^8^ to create survey objects with poststratification weights using the postStratify function and calculate the weighted PMH survey-based prevalence for IHD or stroke, respectively, using the svymeans function. Lastly, we compared the weighted PMH survey-based prevalence estimates against the unweighted estimates to mitigate the impact of non-random likelihood of PMH survey completion.

#### Phenome-wide association scan (PheWAS)

We conducted a PheWAS analysis among participants with EHR to explore clinical phenotypes that may influence the observed prevalence in our study sample and subsamples defined based on the PMH survey completion. We used a Python-based PheTK package to perform PheWAS analyses in the *All of Us* cohort.^9^ Specifically, we used its Phecode module to map and aggregate diagnostic codes (including ICD versions 9-CM, 10, 10-CM, and SNOMED) to 3,409 phecodes using the Phecode X (Extended) ontology (version 1.0).^10^ Participants were considered cases of a given condition if they had two or more qualifying diagnostic codes on separate days. After excluding 1,205 phecodes having less than 100 cases, our PheWAS analysis included 266,472 participants, 2,204 phecodes. To account for potential confounding, we adjusted the PheWAS analysis for age at enrollment, sex assigned at birth, self-reported race and ethnicity, country of birth, age at last clinical encounter documented in EHR, and length of EHR.

#### Data sharing statement

We used data available through the Researcher Workbench for the Registered Tier access (R2022Q4R9), a cloud-based platform where approved researchers can access and analyze data collected from the *All of Us* participants. All analyses were conducted using R Studio version 4.2.3, and Jupyter Notebook with Python, version 3.9, on the Researcher Workbench. The results reported follow the *All of Us* Data and Statistics Dissemination Policy, which does not allow disclosure of results in group counts under 20.

## Results

### Univariate descriptive analysis

**Table 1** presents participant characteristics of the overall sample of participants who released their EHR (N = 266,472), and subsamples stratified by the PMH survey completion: respondents (n = 124,192, 46.6%) and non-respondents (n = 142,280, 53.4%). PMH survey respondents were, on average, significantly older (54.0; standard deviation (SD), 16.6 years vs. 50.4; SD, 16.5 years), more likely to report being female at birth (64.7% vs. 57.4%), and more likely to be married (58.5% vs. 41.0%) than non-respondents. In contrast, non-respondents were more likely to self-identify as Hispanic or Latino (non-respondents: 24.7% vs. respondents: 11.9%) or Black or African American (27.9% vs. 9.8%), and endorsed indicators of lower socioeconomic status, including health insurance coverage (88.2% vs. 94.5%), employment (39.6% vs. 49.3%), and college education or higher (54.8% vs. 82.5%), household income greater than or equal to $50,000 (16.2% vs. 41.5%), and home ownership (32.1% vs. 60.7%). Non-respondents also had higher rates of lifetime cigarette smoking (43.4% vs. 35.7%) but lower rates of lifetime alcohol consumption (82.6% vs. 93.1%). Additionally, non-respondents exhibited lower medical literacy, being approximately three times more likely to lack confidence in filling out medical forms (20.5% vs. 7.0%), have difficulty understanding written medical information (7.9% vs. 2.6%), and need help reading health-related materials (12.2% vs. 4.8%).

**Table 1.** Participant characteristics of the study sample and comparison by Personal Medical History (PMH) survey completion status.

|  | Overall | PMH respondents | PMH non-respondents | *P* |
| --- | --- | --- | --- | --- |
| **Sample size (N (%))** | 266,472 | 124,192 | 142,280 |  |
| ***Demographic characteristics*** |  |  |  |  |
| **Age at enrollment (mean (SD))** | 52.07 (16.7) | 54.00 (16.6) | 50.39 (16.5) | <0.001 |
| **Sex assigned at birth** |  |  |  | <0.001 |
| Female | 161978 (60.8) | 80361 (64.7) | 81617 (57.4) |  |
| Male | 99096 (37.2) | 41204 (33.2) | 57892 (40.7) |  |
| Not male, not female, prefer not to answer, skipped, or no matching concept | 5398 (2.0) | 2627 (2.1) | 2771 (1.9) |  |
| **Birthplace** |  |  |  | <0.001 |
| USA | 221140 (83.0) | 107141 (86.3) | 113999 (80.1) |  |
| Non-USA | 40518 (15.2) | 14607 (11.8) | 25911 (18.2) |  |
| Refused/Skipped | 4814 (1.8) | 2444 (2.0) | 2370 (1.7) |  |
| **Self-reported race and ethnicity** |  |  |  | <0.001 |
| Hispanic or Latino | 49899 (18.7) | 14774 (11.9) | 35125 (24.7) |  |
| Non-Hispanic (NH) Asian | 7147 (2.7) | 3441 (2.8) | 3706 (2.6) |  |
| NH Black or African American | 51849 (19.5) | 12179 (9.8) | 39670 (27.9) |  |
| NH More than one population | 4116 (1.5) | 2073 (1.7) | 2043 (1.4) |  |
| NH White | 141599 (53.1) | 86243 (69.4) | 55356 (38.9) |  |
| Refused/Skipped/None of these | 11862 (4.5) | 5482 (4.4) | 6380 (4.5) |  |
| **Region of residence** |  |  |  | <0.001 |
| Northeast | 80351 (30.2) | 37499 (30.2) | 42852 (30.1) |  |
| Midwest | 59386 (22.3) | 33405 (26.9) | 25981 (18.3) |  |
| South | 45335 (17.0) | 17272 (13.9) | 28063 (19.7) |  |
| West | 66216 (24.8) | 26976 (21.7) | 39240 (27.6) |  |
| Suppressed/Missing | 15184 (5.7) | 9040 (7.3) | 6144 (4.3) |  |
| **Marital status** |  |  |  | <0.001 |
| Married/Living with a partner | 130994 (49.2) | 72591 (58.5) | 58403 (41.0) |  |
| Divorced/Separated/Widowed | 61667 (23.1) | 24755 (19.9) | 36912 (25.9) |  |
| Never Married | 64553 (24.2) | 23534 (18.9) | 41019 (28.8) |  |
| Refused/Skipped | 9258 (3.5) | 3312 (2.7) | 5946 (4.2) |  |
| **Socioeconomic status** |  |  |  |  |
| **Health insurance status** |  |  |  | <0.001 |
| Yes | 242804 (91.1) | 117361 (94.5) | 125443 (88.2) |  |
| No | 14765 (5.5) | 3573 (2.9) | 11192 (7.9) |  |
| Refused/Skipped | 8903 (3.3) | 3258 (2.6) | 5645 (4.0) |  |
| **Employment status** |  |  |  | <0.001 |
| Employed for wages or self-employed | 117547 (44.1) | 61246 (49.3) | 56301 (39.6) |  |
| Not currently employed for wages | 138824 (52.1) | 59490 (47.9) | 79334 (55.8) |  |
| Refused/Skipped | 10101 (3.8) | 3456 (2.8) | 6645 (4.7) |  |
| **Educational attainment** |  |  |  | <0.001 |
| College or higher | 180349 (67.7) | 102398 (82.5) | 77951 (54.8) |  |
| Less than college | 77780 (29.2) | 18736 (15.1) | 59044 (41.5) |  |
| Refused/Skipped | 8343 (3.1) | 3058 (2.5) | 5285 (3.7) |  |
| **Household income** |  |  |  | <0.001 |
| 50k or higher | 74629 (28.0) | 51596 (41.5) | 23033 (16.2) |  |
| Less than 50k | 107975 (40.5) | 38375 (30.9) | 69600 (48.9) |  |
| Refused/Skipped | 83868 (31.5) | 34221 (27.6) | 49647 (34.9) |  |
| **Current homeownership** |  |  |  | <0.001 |
| Own | 121145 (45.5) | 75445 (60.7) | 45700 (32.1) |  |
| Rent | 105247 (39.5) | 36252 (29.2) | 68995 (48.5) |  |
| Other Arrangement | 26449 (9.9) | 8178 (6.6) | 18271 (12.8) |  |
| Refused/Skipped | 13631 (5.1) | 4317 (3.5) | 9314 (6.5) |  |
| **Lifestyle characteristics** |  |  |  |  |
| **Lifetime cigarette smoking** |  |  |  | <0.001 |
| Yes | 106159 (39.8) | 44376 (35.7) | 61783 (43.4) |  |
| No | 154091 (57.8) | 77916 (62.7) | 76175 (53.5) |  |
| Refused/Skipped | 6222 (2.3) | 1900 (1.5) | 4322 (3.0) |  |
| **Lifetime alcohol consumption** |  |  |  | <0.001 |
| Yes | 233146 (87.5) | 115650 (93.1) | 117496 (82.6) |  |
| No | 29566 (11.1) | 7816 (6.3) | 21750 (15.3) |  |
| Refused/Skipped | 3760 (1.4) | 726 (0.6) | 3034 (2.1) |  |
| **Medical literacy characteristics** |  |  |  |  |
| **How confident are you in filling out medical forms by yourself?** |  |  |  | <0.001 |
| Not At All/A Little Bit/Somewhat | 37834 (14.2) | 8704 (7.0) | 29130 (20.5) |  |
| Quite A Bit/Extremely | 224548 (84.3) | 114698 (92.4) | 109850 (77.2) |  |
| Skipped | 4090 (1.5) | 790 (0.6) | 3300 (2.3) |  |
| **How often do you have problems learning about your medical condition because of difficulty understanding written information?** |  |  |  | <0.001 |
| Never/Occasionally/Sometimes | 245931 (92.3) | 119721 (96.4) | 126210 (88.7) |  |
| Often/Always | 14404 (5.4) | 3182 (2.6) | 11222 (7.9) |  |
| Skipped | 6137 (2.3) | 1289 (1.0) | 4848 (3.4) |  |
| **How often do you have someone help you read health-related materials?** |  |  |  | <0.001 |
| Never/Occasionally/Sometimes | 238463 (89.5) | 117332 (94.5) | 121131 (85.1) |  |
| Often/Always | 23361 (8.8) | 5986 (4.8) | 17375 (12.2) |  |
| Skipped | 4648 (1.7) | 874 (0.7) | 3774 (2.7) |  |

### Multivariable logistic regression analysis

As summarized in **Figure 1** and **eTable 2**, older age at enrollment was associated with a 9% increase in the odds of completing the PMH survey for each standard deviation increase in age (OR, 1.09; 95% CI, 1.08-1.11). Compared to participants reporting female at birth, those reporting male sex at birth had over a 30% decrease in the odds of completing the PMH survey (OR, 0.68; 95% CI, 0.67-0.70). Participants who reported neither female nor male sex at birth had a 20% increase in the odds of PMH survey completion (OR, 1.20; 95% CI, 1.11-1.29). Participants identifying as Black or African American were significantly less likely than non-Hispanic White participants to complete the PMH survey, with up to 63% reduction in odds (OR, 0.37; 95% CI, 0.36-0.38). Similarly, Hispanic or Latino participants were 50% less likely to complete the PMH survey (OR, 0.50; 95% CI, 0.48-0.51).

**Figure 1.**
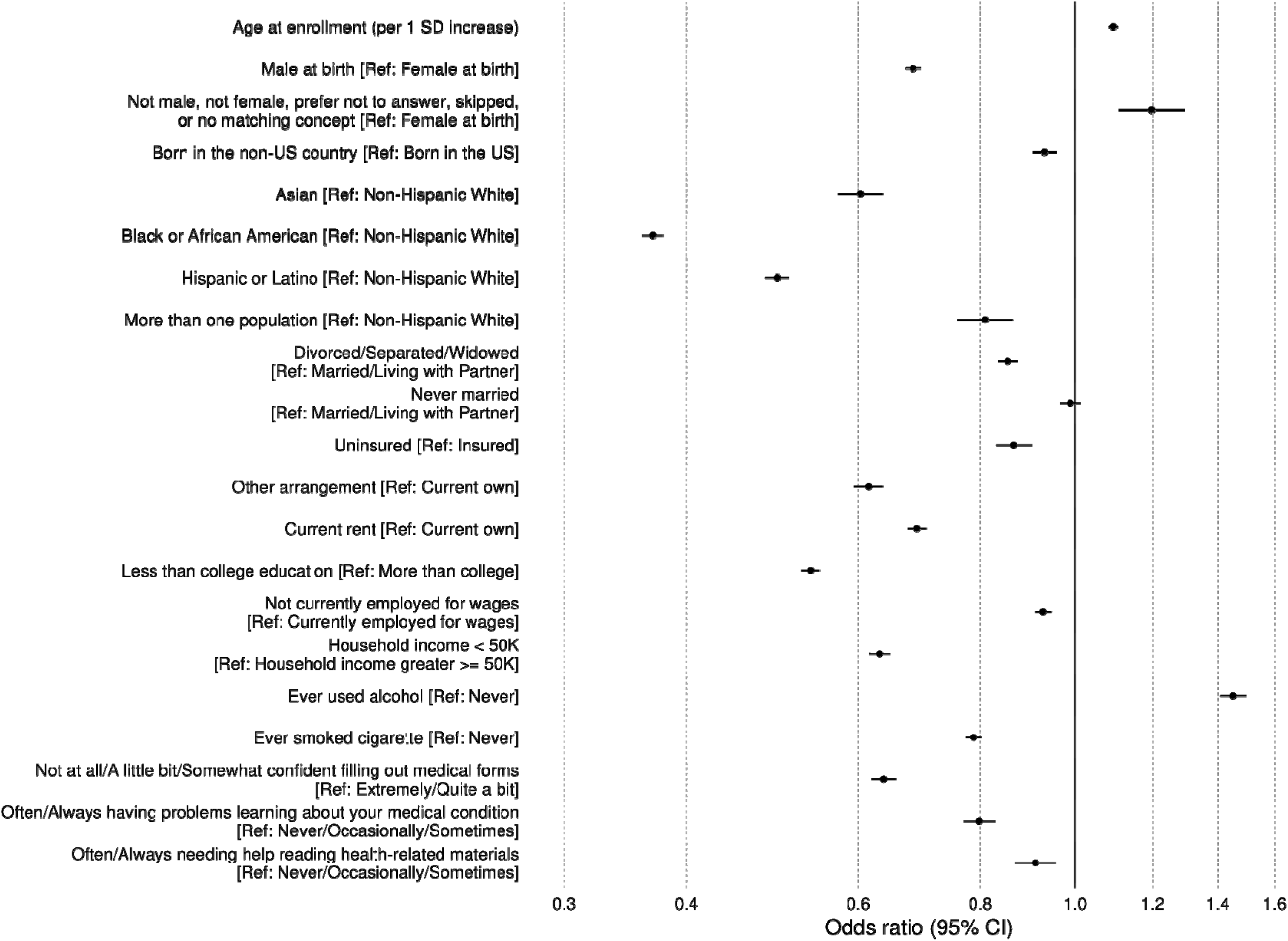
Multivariate logistic regression analysis predicting the PMH survey completion among participants with EHR (N=266,472).

Characteristics indicative of lower socioeconomic status generally corresponded to decreased odds of PMH survey completion. For instance, renters were nearly 30% less likely to complete the survey compared to homeowners (OR, 0.69; 95% CI, 0.67-0.70), and those with less than a college education had a nearly 50% lower likelihood of completing the survey (OR, 0.54; 95% CI, 0.52-0.55). Participants with an annual household income under $50,000 were approximately 40% less likely to complete the survey (OR, 0.63; 95% CI, 0.62-0.65). Interestingly, a lifetime history of alcohol consumption was associated with a 45% increase in the odds of survey completion (OR, 1.45; 95% CI, 1.41-1.50), while a lifetime history of cigarette smoking was associated with a 20% decrease in odds (OR, 0.79; 95% CI, 0.77-0.80). Lastly, individuals reporting lower confidence in completing medical forms, difficulties in understanding medical information, or needing assistance with health-related materials were significantly less likely to complete the PMH survey, with a reduction in odds up to 36% (OR, 0.64; 95% CI, 0.62-0.66 for lacking confidence in filling out medical forms).

### Prevalence of IHD

The EHR-based prevalence of IHD in the overall sample of 266,472 participants was 6.4% (95% CI, 6.3-6.5). When stratified by PMH survey completion status, the EHR-based prevalence was higher among non-respondents at 7.2% (95% CI, 7.0-7.3) compared to 5.6% (95% CI, 5.4-5.7) among respondents (see **Figure 2** and **eTable 3**). Among 124,192 participants who additionally completed the PMH survey, the PMH-based prevalence of IHD was 5.9% (95% CI, 5.7-6.0), which remained nearly unchanged after poststratification weighting for household income and educational attainment (see **Figure 3** and **eTable 3**).

**Figure 2.**
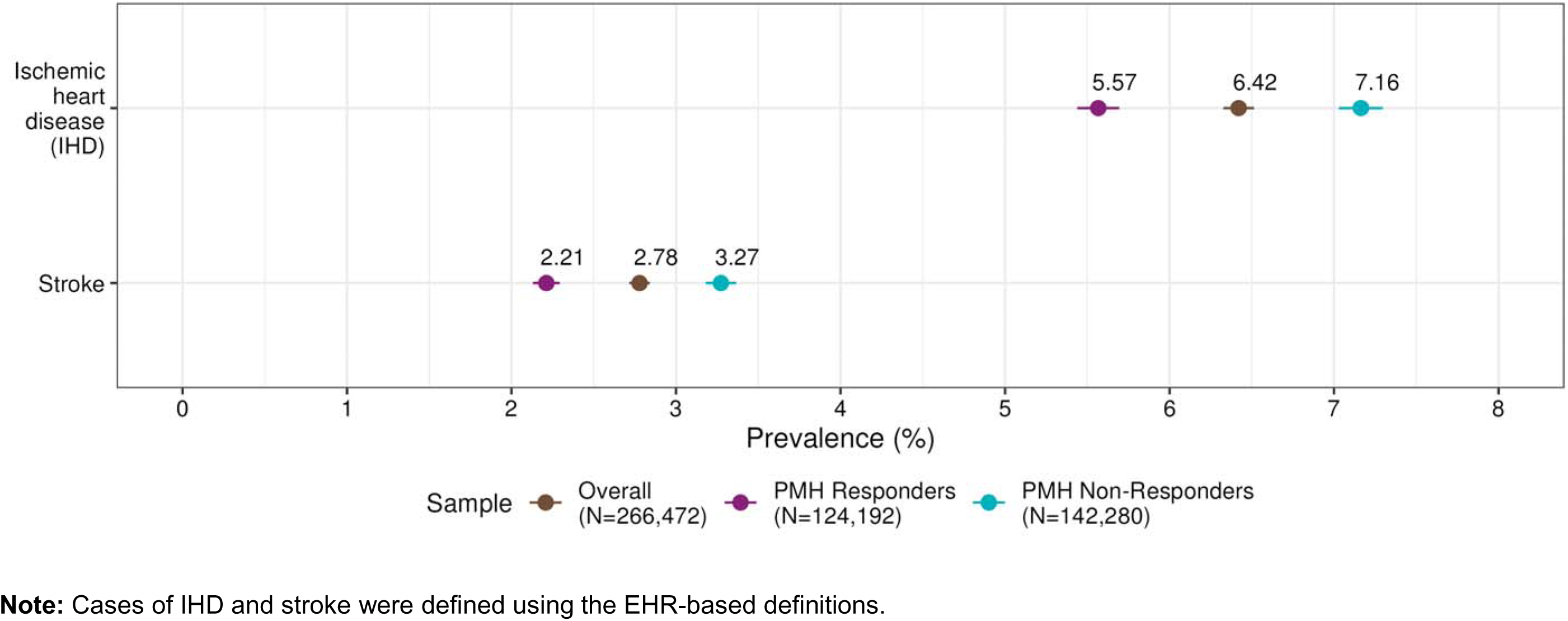
EHR-based prevalence estimates for ischemic heart disease (IHD) and stroke, stratified by PMH survey completion status.

**Figure 3.**
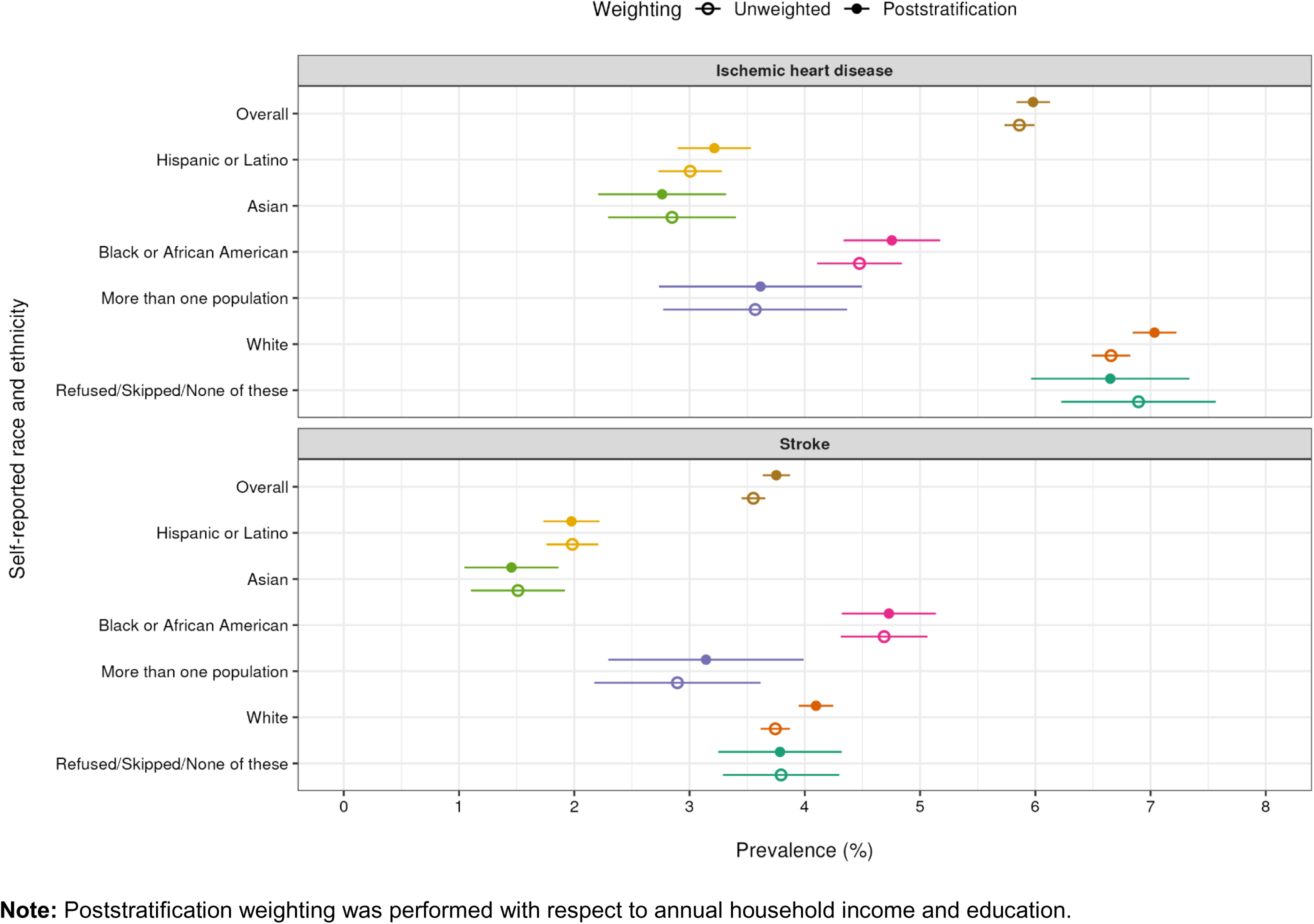
PMH survey-based prevalence estimates of IHD and stroke with or without poststratification weighting.

There were substantial variations in the PMH-based prevalence of IHD when stratified by self-reported race and ethnicity (see **Figure 3** and **eTable 4**). In the unweighted estimation, we found the highest prevalence of IHD among participants who either refused, skipped, or selected “none of these” for their race and ethnicity (6.9%; 95% CI, 6.2-7.6), followed by NH White participants (6.7%; 95% CI, 6.5-6.8), NH Black or African American participants (4.5%; 95% CI, 4.1-4.8), and the lowest among NH Asian participants (2.8%; 95% CI, 2.3-3.4). After applying poststratification weights, we found the highest prevalence among NH White participants (7.0%; 95% CI, 6.8-7.2) and the lowest among non-Hispanic Asian participants (2.8%; 95% CI, 2.2-3.3).

### Prevalence of stroke

The EHR-based prevalence of stroke in the overall sample was 2.8% (95% CI, 2.7-2.8). Similar to IHD, the prevalence was higher among non-respondents at 3.3% (95% CI, 3.2-3.4) and lower among respondents of the PMH survey at 2.2% (95% CI, 2.1-2.3) (see **Figure 2** and **eTable 3)**. Among those who additionally completed the PMH survey, the PMH-based prevalence of stroke was 3.6% (95% CI, 3.5-3.7), increasing slightly with poststratification weighting to 3.8% (95% CI, 3.6-3.9) (see **Figure 3** and **eTable 3**).

There were notable variations in the PMH-based prevalence of stroke by self-reported race and ethnicity (see **Figure 3** and **eTable 4**). Participants who self-identified as NH Black or African American had the highest prevalence of stroke (4.7%; 95% CI, 4.3-5.1), followed by those refused, skipped, or selected “none of these” for their race and ethnicity (3.8%; 95% CI, 3.3-4.3) and non-Hispanic White participants (3.7%; 95% CI, 3.6-3.9). While poststratification weighting had a minimal impact on the PMH-based prevalence overall, it significantly increased the prevalence from 3.7% (95% CI, 3.6-3.9) to 4.1% (95% CI, 3.9-4.2) among NH White participants.

### PheWAS

There were diagnostic codes associated with a significantly *increased* likelihood of PMH survey completion among participants who released the EHR (see **Figure 4a**). These codes corresponded to less critical health conditions indicative of access to routine healthcare, including dermatological conditions (e.g., benign neoplasm of the skin, melanocytic nevi, seborrheic keratosis, acne), sensory conditions (e.g. myopia, astigmatism, hearing loss), and respiratory conditions (e.g., allergic rhinitis, rhinitis and nasal congestion).

**Figure 4.**
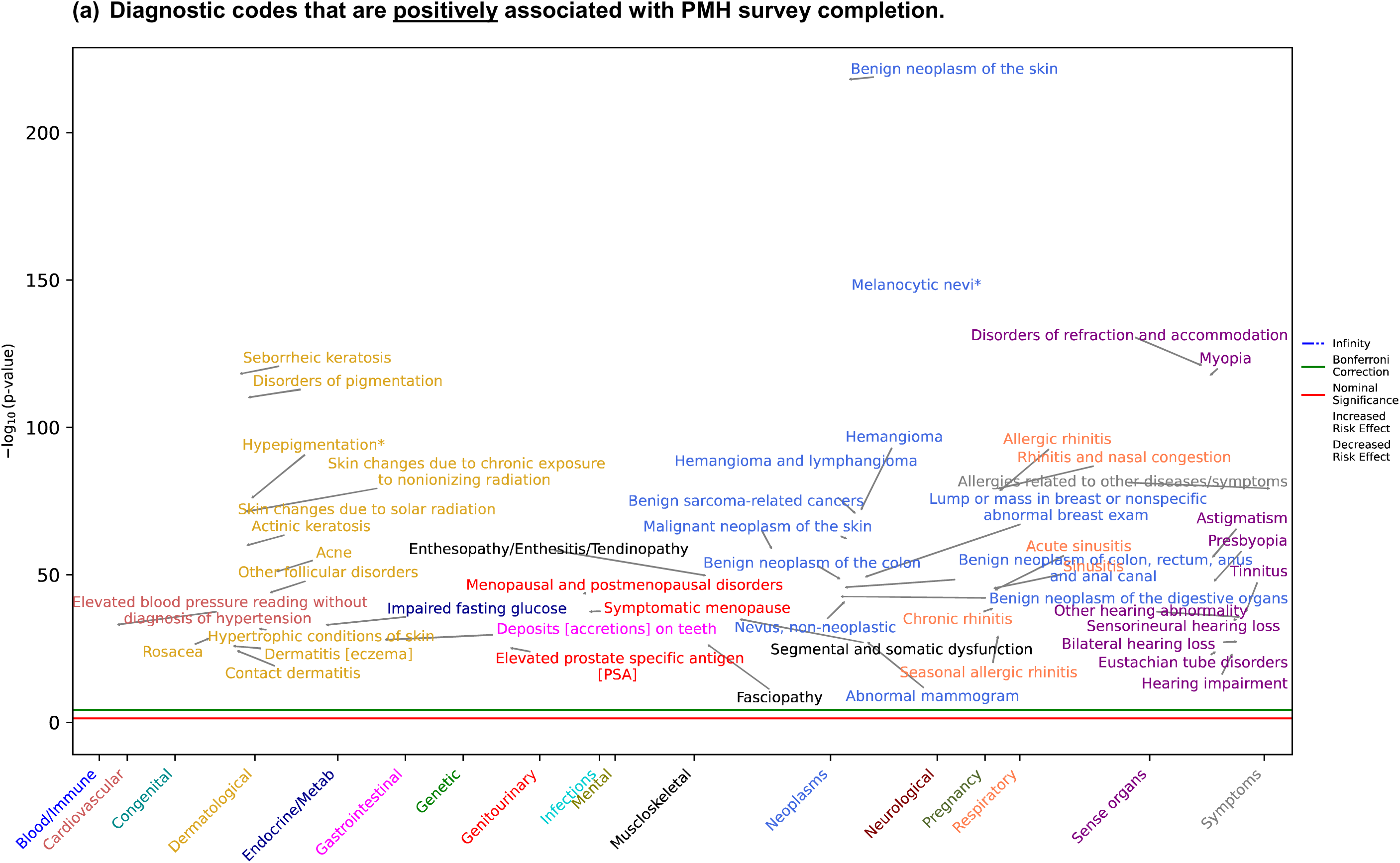
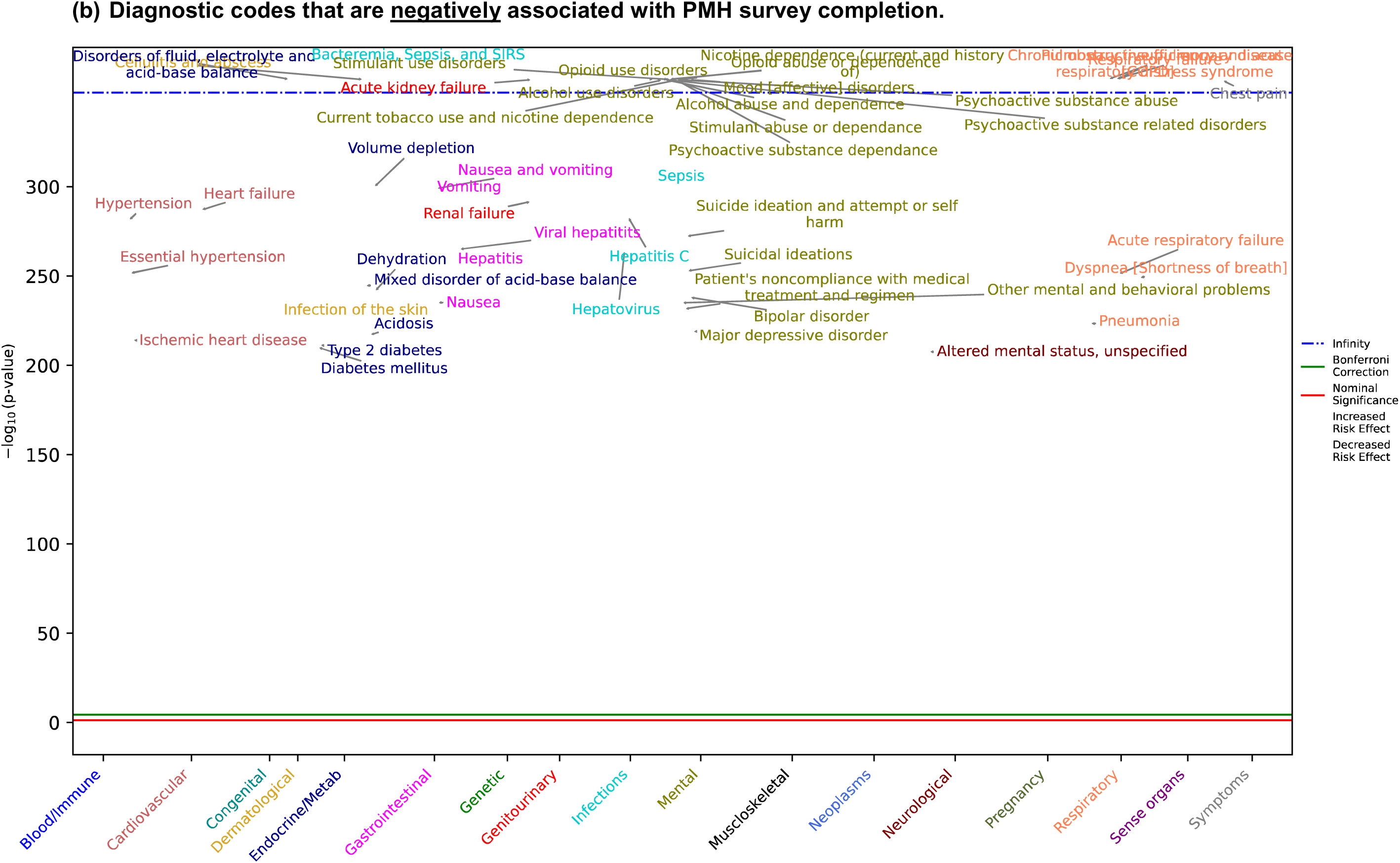
Manhattan plot summarizing results from the phenome-wide association study (PheWAS) of PMH survey completion.

Conversely, there were diagnostic codes associated with a significantly *decreased* likelihood of PMH survey completion (see **Figure 4b**). They included critical conditions requiring specialized care, such as psychiatric conditions (e.g., substance use, suicidal behavior, bipolar disorder, major depressive disorder), endocrine and genitourinary conditions (e.g., volume depletion, type 2 diabetes, kidney failure), and most importantly, cardiovascular conditions (e.g., heart failure, essential hypertension, ischemic heart disease).

## Discussion

In the present study, we investigated the impact of participation bias on prevalence estimation using EHR and the PMH survey, focusing on IHD and stroke as case examples. We first estimated the prevalence of these conditions in the overall sample of 266,472 participants with EHR and compared prevalence in the subsamples stratified by PMH survey completion. The EHR-based prevalence estimates for IHD and stroke among participants with EHR were 6.4% and 2.8%, respectively. These estimates align closely with recent population-based estimates from the Behavioral Risk Factor Surveillance System, which reported prevalence estimates of 6.0%^11^ for IHD and 2.9%^12^ for stroke. The *All of Us* Program is not designed to represent the US general population and is typically not considered an ideal data source for estimating disease prevalence.^13^ However, our findings suggest that its current EHR cohort may resemble the US general population for IHD and stroke, offering new opportunities for future research.

We subsequently estimated the EHR-based prevalence of IHD and stroke, stratified by PMH survey completion. The EHR-based prevalence estimates were consistently lower among PMH survey respondents but higher among non-respondents. More specifically, PMH survey respondents were less diverse and had higher education, income, and medical literacy than non-respondents, consistent with the well-established “healthy volunteer” bias.^14^ To address the skewed distribution of participant characteristics in PMH survey responses, we applied poststratification weights adjusting for annual household income and educational attainment. For both IHD and stroke, the weighted PMH-survey based prevalence estimates were higher than the unweighted estimates, accounting for the differences between the overall study sample and PMH respondents by household income and education. Additionally, we found substantial variations in the prevalence of IHD and stroke by self-reported race and ethnicity. Notably, the effect of poststratification weighting was particularly pronounced for stroke and among participants self-identifying as NH White. The weighted PMH survey-based prevalence was significantly and consistently higher than the unweighted prevalence among NH White participants. These results underscore the importance of bias mitigation when using voluntary follow-up modules to ensure accurate estimation of disease burden.

Our findings contribute to the growing body of literature emphasizing the importance of adjusting for the non-random likelihood of participation in follow-up modules in longitudinal cohort studies. Previous studies, such as those conducted in the Avon Longitudinal Study of Parents and Children (ALSPAC)^15,16^ and the UK Biobank,^6,17^ have highlighted this issue. In the context of the *All of Us* Research Program, our team previously reported on the impact of non-random participation in its COVID-19 Participant Experience (COPE) survey administered during the pandemic.^18^ Consistent with the findings from the ALSPAC and UK Biobank, we observed that COPE survey respondents were more likely to be female, non-Hispanic White, older, have higher socioeconomic status and lower mental health burden compared to the underlying *All of Us* cohort.^18^ In our previous studies using the COPE surveys, we applied a causal inference method called inverse probability weighting^19^ to mitigate bias when estimating of hypothesized causal relationships.^18,20^

In the current study, we applied poststratification weighting to mitigate healthy volunteer bias in the context of disease prevalence estimation. Calibration and propensity score weighting are two most common approaches to post-survey adjustment.^21^ The simplest form of calibration is poststratification, which divides the nonprobability samples (i.e., PMH survey respondents) into mutually exclusive cells and weights them to match the corresponding proportion in the target population (i.e., EHR cohort).^22^ After applying poststratification weights for annual household income and education, we found that the EHR-based prevalence of IHD and stroke was underestimated when non-random sampling was not accounted for. This finding underscores the importance of addressing participation bias in voluntary follow-up modules to ensure accurate estimation of disease burdens and associations in target populations.

In addition to sociodemographic and medical literacy characteristics, we conducted a PheWAS analysis to investigate the clinical characteristics that further influence the likelihood of the PMH survey completion. Our results showed that diagnostic codes indicative of access to routine healthcare (e.g., benign neoplasm of the skin, myopia, allergic rhinitis, astigmatism) were associated with an *increased* likelihood of PMH survey completion. Conversely, diagnostic codes indicative of critical and debilitating conditions, including those strongly associated with IHD and stroke (e.g., hypertension, heart failure), were associated with a *decreased* likelihood of PMH survey completion, potentially resulting in an incomplete capture of important health outcomes among PMH survey respondents. These findings provide a nuanced explanation for the potential differences in access to routine healthcare and underlying disease burden between PMH survey respondents and non-respondents. Additionally, they highlight the broader complexity involved in interpreting the observed differences between the unweighted and weighted prevalence estimates. Future studies may consider incorporating a more comprehensive set of characteristics that differentiate volunteers from the target population as it continues to expand its follow-up modules.^23^

Taken together, our findings demonstrate that prevalence estimation can be sensitive to the sample characteristics, which are fundamentally shaped by recruitment and data collection strategies. Despite the *All of Us* Research Program being a volunteer-based cohort, its EHR-based prevalence estimates of IHD and stroke closely resembled national estimates. This may be due to the Program’s efforts to oversample participants from historically underrepresented contexts in biomedical studies (e.g., lower education, income, racial and ethnic diversity). We found a slight underestimation of EHR-based prevalence when restricted to participants who additionally completed the PMH survey, possibly impacted by the reduced representation of underprivileged sociodemographic contexts relative to the overall study sample. Therefore, when working with voluntary follow-up modules in longitudinal cohort studies, it is important for researchers to clearly define the target population and evaluate how sample characteristics vary across data modalities and assessment timepoints, accurately representing the disease burden.

## Conclusion

In conclusion, the present study demonstrates that disease prevalence estimation can be biased if non-random sampling is not adequately addressed in follow-up modules relying on active participant engagement in volunteer-based cohorts, such as the *All of Us* Research Program.

## Supporting information

eMethods 1

eMethods 2

eMethods 3

eTable 1

eTable 2

eTable 3

eTable 4

## Data Availability

Access to the Registered and Controlled Tier data is restricted to US-based researchers whose institutions have signed a Data Use and Registration Agreement (DURA) and who have verified their identity.

https://workbench.researchallofus.org/

## Acknowledgements

This project was supported by funding through the National Institutes of Health, Office of the Director (OT2OD026553). In addition, the *All of Us* Research Program would not be possible without the contributions made by its participants.

## References

1. Multiple Cause of Death Data on CDC WONDER. https://wonder.cdc.gov/mcd.html.

2. Wadhera, R. K. et al. Racial and ethnic disparities in heart and cerebrovascular disease deaths during the COVID-19 pandemic in the United States. Circulation 143, 2346–2354 (2021).

3. Kyalwazi, A. N. et al. Disparities in cardiovascular mortality between Black and White adults in the United States, 1999 to 2019. Circulation 146, 211–228 (2022).

4. Ilori, T. O. et al. Approach to high volume enrollment in clinical research: Experiences from an All of Us Research Program site. Clin. Transl. Sci. 13, 685–692 (2020).

5. All of Us Research Program Investigators et al. The “All of Us” Research Program. N. Engl. J. Med. 381, 668–676 (2019).

6. Tyrrell, J. et al. Genetic predictors of participation in optional components of UK Biobank. Nat. Commun. 12, 886 (2021).

7. Acosta, J. N. et al. Cardiovascular health disparities in racial and other underrepresented groups: Initial results from the All of Us research program. J. Am. Heart Assoc. 10, e021724 (2021).

8. Lumley T, Gao P, Schneider B. Survey: Analysis of Complex Survey Samples. (The R Foundation, 2024). doi:10.32614/cran.package.survey.

9. Tran, T. C. et al. PheWAS analysis on large-scale biobank data with PheTK. medRxiv (2024) doi:10.1101/2024.02.12.24302720.

10. Shuey, M. M. et al. Next-generation phenotyping: introducing phecodeX for enhanced discovery research in medical phenomics. Bioinformatics 39, btad655 (2023).

11. Lee, Y.-T. H. et al. Prevalence and trends of coronary heart disease in the United States, 2011 to 2018. JAMA Cardiol. 7, 459–462 (2022).

12. Imoisili, O., Chung, A., Tong, X., Hayes, D. & Loustalot, F. Prevalence of stroke — Behavioral Risk Factor Surveillance System, United States, 2011–2022. Morbidity and Mortality Weekly Report 73, 449–455 (2024).

13. Kathiresan, N. et al. Representation of race and ethnicity in the contemporary US health cohort all of Us Research Program. JAMA Cardiol. 8, 859–864 (2023).

14. Delgado-Rodríguez, M. & Llorca, J. Bias. J. Epidemiol. Community Health 58, 635–641 (2004).

15. Martin, J. et al. Association of genetic risk for schizophrenia with nonparticipation over time in a population-based cohort study. Am. J. Epidemiol. 183, 1149–1158 (2016).

16. Taylor, A. E. et al. Exploring the association of genetic factors with participation in the Avon Longitudinal Study of Parents and Children. Int. J. Epidemiol. 47, 1207–1216 (2018).

17. Adams, M. J. et al. Factors associated with sharing e-mail information and mental health survey participation in large population cohorts. Int. J. Epidemiol. 49, 410–421 (2020).

18. Lee, Y. H. et al. Association of everyday discrimination with depressive symptoms and suicidal ideation during the COVID-19 pandemic in the all of Us Research Program. JAMA Psychiatry 79, 898–906 (2022).

19. Hernán, M. A. & Robins, J. M. Estimating causal effects from epidemiological data. J. Epidemiol. Community Health 60, 578–586 (2006).

20. Choi, K. W. et al. Social support and depression during a global crisis. Nat. Ment. Health 1, 428–435 (2023).

21. Earp, M., Mitchell, M., Kott, P., Kreuter, F. & Porter, E. Nonresponse Bias Adjustment in Establishment Surveys: A Comparison of Weighting Methods using the Agricultural Resource Management Survey (ARMS). Bureau of Labor Statistics https://www.bls.gov/osmr/research-papers/2012/st120240.htm (2019).

22. Mercer, A. W., Kreuter, F., Keeter, S. & Stuart, E. A. Theory and practice in nonprobability surveys. Public Opin. Q. 81, 250–271 (2017).

23. Downes, M. et al. Multilevel regression and poststratification: A modeling approach to estimating population quantities from highly selected survey samples. Am. J. Epidemiol. 187, 1780–1790 (2018).

