## Supplementary material for "Impact of Participation Bias on Disease Prevalence Estimation in the *All of Us* Research Program: A Case Study of Ischemic Heart Disease and Stroke": eMethods 1

**eMethods 1.** Participant recruitment and data collection in the NIH *All of Us* Research Program.

Details of the design and scientific rationale of the *All of Us* Research Program have been described previously.^1^ Briefly, beginning in May 2018, participants aged ≥18 years were enrolled from recruitment sites across all 50 states and will continue until ≥1 million participants are enrolled ^1^. The *All of Us* Research Program is designed to recruit people who are underrepresented in biomedical research with the goal of enrolling its cohort from populations that are more than 75% underrepresented in terms of demographics, geographic location, and other characteristics, with at least 45% of participants coming from racial and ethnic groups that are underrepresented in research.^1^

Information on the sites from which participants are recruited has been described.^1,2^ Briefly, recruitment sites for *All of Us* were selected via an NIH submission and review process. The number of recruitment sites is evolving, and as of this writing, *All of Us* participants were enrolled at regional medical centers, Federally Qualified Health Centers, Veterans Health Administration sites, and “direct volunteer” sites that can provide access for people who are not patients in a healthcare organization (a designated health clinic, blood bank, laboratory, or other facility). Participants in *All of Us* enroll digitally and provide informed consent to participate in the program through the website (https://www.joinallofus.org), via a smartphone application, or through one of the participating recruitment sites. After a person 1) consents to participate, 2) provides authorization to share EHR data, and 3) completes the initial baseline survey of demographic information, the participant becomes eligible for in-person visits to have physical measurements and biospecimens collected at one of the *All of Us* recruitment sites. Our analysis of de-identified data was classified as research that did not involve human subjects by the *All of Us* institutional review board.

Further details of the surveys and data collection methods are available in the Survey Explorer found in the *All of Us* Research Hub (https://www.researchallofus.org/), a website designed to support researchers. Three currently available data modalities (survey, physical measurements, and EHR) are mapped to the Observational Health and Medicines Outcomes Partnership (OMOP) common data model, version 5.2, maintained by the Observational Health and Data Sciences Initiative collaborative. To protect participant privacy, a series of data transformations were applied. These included data suppression of codes with a high risk of identification, such as military status; generalization of categories, including age, sex at birth, gender identity, sexual orientation, and race or ethnicity; and date shifting by a random (less than 1 year) number of days, implemented consistently across each participant record.

**References**

1.	All of Us Research Program Investigators *et al.* The “All of Us” Research Program. *N. Engl. J. Med.* **381**, 668–676 (2019).

2.	Ramirez, A. H. *et al.* The All of Us Research Program: Data quality, utility, and diversity. *Patterns (N. Y.)* **3**, 100570 (2022).
