## Supplementary material for "Impact of Participation Bias on Disease Prevalence Estimation in the *All of Us* Research Program: A Case Study of Ischemic Heart Disease and Stroke": eMethods 2

**eMethods 2.** The Heart and Blood conditions section of the Personal Medical History survey.

*All of Us* Research Program

Participant Provided Information (PPI)

Version: October 12, 2018

**Personal Medical History**

This survey asks questions about your Personal Medical History. This is to better understand how it may affect health. To ensure your privacy, your name will be separated from your answers before they are shared with researchers.

It will take about 10-15 minutes to answer these questions. Please answer each question as honestly as possible. Some of the questions may be sensitive. You can choose not to answer. There are no right or wrong answers to any of the questions. It is important that you answer as many questions as you can. We are looking for your own answers, not what you think your doctors, family, or friends want you to say.

Do not feel like you have to spend a long time on each question. The first answer that comes to you is usually the best one. If you are not sure how to answer a question, choose the best answer from the options given. If you are unsure of what a condition is, hover over the ‘i’ icon to its right for more information.

Heart and blood conditions: Has a doctor or health care provider ever told you that you have…?

(select all that apply)

•	Anemia

•	Atrial fibrillation (Afib) or Atrial flutter

•	Bleeding disorder

•	Congestive heart failure

•	**Coronary artery/coronary heart disease**

•	**Heart attack**

•	Heart valve disease

•	High cholesterol

•	Hypertension (high blood pressure)

•	Peripheral vascular disease

•	Pulmonary embolism or deep vein thrombosis

•	Sickle cell disease

•	**Stroke**

•	**Transient ischemic attacks (TIAs or mini-strokes)**

•	Other heart or blood condition

•	I have no heart or blood condition

*Branching logic*: If any of the above conditions are selected, display the following associated questions:

- **Coronary artery/coronary heart disease:**
  - Are you still seeing a doctor or health care provider for this condition? Yes/No
  - About how old were you when you were first told you had this condition?
    - Child (0-11)
    - Adolescent (12-17)
    - Adult (18-64)
    - Older adult (65-74)
    - Elderly (75+)
  - Are you currently prescribed medications and/or receiving treatment for this condition? Yes/No

- **Heart attack:**
  - Are you still seeing a doctor or health care provider for this condition? Yes/No
  - About how old were you when you were first told you had this condition?
    - Child (0-11)
    - Adolescent (12-17)
    - Adult (18-64)
    - Older adult (65-74)
    - Elderly (75+)
  - Are you currently prescribed medications and/or receiving treatment for this condition? Yes/No
- **Stroke:**
  - Are you still seeing a doctor or health care provider for this condition? Yes/No
  - About how old were you when you were first told you had this condition?
    - Child (0-11)
    - Adolescent (12-17)
    - Adult (18-64)
    - Older adult (65-74)
    - Elderly (75+)
  - Are you currently prescribed medications and/or receiving treatment for this condition? Yes/No

- **Transient ischemic attacks (TIAs or mini-strokes):**
- Are you still seeing a doctor or health care provider for this condition? Yes/No
- About how old were you when you were first told you had this condition?
- Child (0-11)
- Adolescent (12-17)
- Adult (18-64)
- Older adult (65-74)
- Elderly (75+)
- Are you currently prescribed medications and/or receiving treatment for this condition? Yes/No
