## Supplementary material for "Impact of Participation Bias on Disease Prevalence Estimation in the *All of Us* Research Program: A Case Study of Ischemic Heart Disease and Stroke": eMethods 3

**eMethods 3.** Survey questions used to measure health literacy from the Overall Health survey.

*All of Us* Research Program

Participant Provided Information (PPI)

Version: May 31, 2018

**Overall Health**

This survey asks questions about your overall health. Your privacy is very important to us. Your answers will only be shared with approved researchers after we have removed your name. It takes about 5-10 minutes to answer these questions. Please answer each question as honestly as possible. There are no right or wrong answers to any of the questions. It is important that you answer as many questions as you can. We are looking for your own answers, and not what you think your doctors, family, or friends want you to say.

Don't feel like you have to spend a long time over each question. The first answer that comes to you is usually the best one. If you aren't sure how to answer a question, choose the best answer from the options given.

**How confident are you filling out medical forms by yourself?**

- Extremely
- Quite a bit
- Somewhat
- A little bit
- Not at all

**How often do you have someone help you read health-related materials?**

- Always
- Often
- Sometimes
- Occasionally
- Never

**How often do you have problems learning about your medical condition because of difficulty understanding written information?**

- Always
- Often
- Sometimes
- Occasionally
- Never

Source:

1. Brief Health Literacy Screen (BHLS)
