## Supplementary material for "Impact of Participation Bias on Disease Prevalence Estimation in the *All of Us* Research Program: A Case Study of Ischemic Heart Disease and Stroke": eTable 1

**eTable 1.** A complete list of diagnostic and procedure codes included in the EHR-based definitions of ischemic heart disease (IHD) and stroke.

| **ICD-9 CM Condition Codes** | **Code** | **Description** |
| --- | --- | --- |
|  | 410 | Acute myocardial infarction |
|  | 411 | Other acute and subacute forms of ischemic heart disease |
|  | 411.1 | Unstable Angina |
|  | 411.9 | Other acute and subacute forms of ischemic heart disease |
|  | 412 | Old myocardial infarction |
|  | 413 | Angina pectoris |
|  | 414 | Other forms of chronic ischemic heart disease |
|  | 414 | Coronary atherosclerosis |
|  | 414.2 | Chronic total occlusion of coronary artery |
|  | 414.3 | Coronary atherosclerosis due to lipid rich plaque |
|  | 414.4 | Coronary atherosclerosis due to calcified coronary lesion |
|  | 414.8 | Other specified forms of chronic ischemic heart disease |
|  | 414.9 | Chronic ischemic heart disease unspecified |
|  | 429.7 | Certain sequelae of myocardial infarction, not elsewhere classified |
|  | V45.81 | Aortocoronary bypass status |
|  | V45.82 | Percutaneous transluminal coronary angioplasty status |
| **ICD-10 CM Condition Codes** | I20 | Unstable Angina |
|  | I21 | Acute myocardial infarction |
|  | I21.0 | ST elevation (STEMI) myocardial infarction of anterior wall |
|  | I21.01 | ST elevation (STEMI) myocardial infarction involving left main coronary artery |
|  | I21.02 | ST elevation (STEMI) myocardial infarction involving left anterior descending coronary artery |
|  | I21.09 | ST elevation (STEMI) myocardial infarction involving other coronary artery of anterior wall |
|  | I21.1 | ST elevation (STEMI) myocardial infarction of inferior wall |
|  | I21.11 | ST elevation (STEMI) myocardial infarction involving right coronary artery |
|  | I21.19 | ST elevation (STEMI) myocardial infarction involving other coronary artery of inferior wall |
|  | I21.2 | ST elevation (STEMI) myocardial infarction of other sites |
|  | I21.21 | ST elevation (STEMI) myocardial infarction involving left circumflex coronary artery |
|  | I21.29 | ST elevation (STEMI) myocardial infarction involving other sites |
|  | I21.3 | ST elevation (STEMI) myocardial infarction of unspecified site |
|  | I21.4 | Non-ST elevation (NSTEMI) myocardial infarction |
|  | I21.9 | Acute myocardial infarction, unspecified |
|  | I21.A | Other type of myocardial infarction |
|  | I21.A1 | Myocardial infarction type 2 |
|  | I21.A9 | Other myocardial infarction type |
|  | I22 | Subsequent ST elevation (STEMI) and non-ST elevation (NSTEMI) myocardial infarction |
|  | I22.0 | Subsequent ST elevation (STEMI) myocardial infarction of anterior wall |
|  | I22.1 | Subsequent ST elevation (STEMI) myocardial infarction of inferior wall |
|  | I22.2 | Subsequent non-ST elevation (NSTEMI) myocardial infarction |
|  | I22.8 | Subsequent ST elevation (STEMI) myocardial infarction of other sites |
|  | I22.9 | Subsequent ST elevation (STEMI) myocardial infarction of unspecified site |
|  | I23 | Certain current complications following ST elevation (STEMI) and non-ST elevation (NSTEMI) myocardial infarction (within the 28 day period) |
|  | I23.0 | Hemopericardium as current complication following acute myocardial infarction |
|  | I23.1 | Atrial septal defect as current complication following acute myocardial infarction |
|  | I23.2 | Ventricular septal defect as current complication following acute myocardial infarction |
|  | I23.3 | Rupture of cardiac wall without hemopericardium as current complication following acute myocardial infarction |
|  | I23.5 | Rupture of papillary muscle as current complication following acute myocardial infarction |
|  | I23.6 | Thrombosis of atrium, auricular appendage, and ventricle as current complications following acute myocardial infarction |
|  | I23.7 | Postinfarction angina |
|  | I23.8 | Other current complications following acute myocardial infarction |
|  | I24 | Other acute ischemic heart diseases |
|  | I24.0 | Acute coronary thrombosis not resulting in myocardial infarction |
|  | I24.1 | Dressler's syndrome |
|  | I24.8 | Other forms of acute ischemic heart disease |
|  | I24.9 | Acute ischemic heart disease, unspecified |
|  | I25 | Chronic Ischemic Heart Disease |
|  | I25.1 | Atherosclerotic heart disease of native coronary artery |
|  | I25.110 | Atherosclerotic heart disease of native coronary artery with unstable angina pectoris |
|  | I25.111 | Atherosclerotic heart disease of native coronary artery with angina pectoris with documented spasm |
|  | I25.118 | Atherosclerotic heart disease of native coronary artery with other forms of angina pectoris |
|  | I25.119 | Atherosclerotic heart disease of native coronary artery with unspecified angina pectoris |
|  | I25.2 | Old myocardial infarction |
|  | I25.5 | Ischemic cardiomyopathy |
|  | I25.6 | Silent myocardial ischemia |
|  | I25.700 | Atherosclerosis of coronary artery bypass graft(s), unspecified, with unstable angina pectoris |
|  | I25.701 | Atherosclerosis of coronary artery bypass graft(s), unspecified, with angina pectoris with documented spasm |
|  | I25.708 | Atherosclerosis of coronary artery bypass graft(s), unspecified, with other forms of angina pectoris |
|  | I25.709 | Atherosclerosis of coronary artery bypass graft(s), unspecified, with unspecified angina pectoris |
|  | I25.710 | Atherosclerosis of autologous vein coronary artery bypass graft(s) with unstable angina pectoris |
|  | I25.711 | Atherosclerosis of autologous vein coronary artery bypass graft(s) with angina pectoris with documented spasm |
|  | I25.718 | Atherosclerosis of autologous vein coronary artery bypass graft(s) with other forms of angina pectoris |
|  | I25.719 | Atherosclerosis of autologous vein coronary artery bypass graft(s) with unspecified angina pectoris |
|  | I25.720 | Atherosclerosis of autologous artery coronary artery bypass graft(s) with unstable angina pectoris |
|  | I25.721 | Athscl autologous artery CABG w ang pctrs w documented spasm |
|  | I25.728 | Atherosclerosis of autologous artery coronary artery bypass graft(s) with other forms of angina pectoris |
|  | I25.729 | Atherosclerosis of autologous artery coronary artery bypass graft(s) with unspecified angina pectoris |
|  | I25.730 | Atherosclerosis of nonautologous biological coronary artery bypass graft(s) with unstable angina pectoris |
|  | I25.731 | Athscl of nonautologous biological CABG w ang pctrs w documented spasm |
|  | I25.738 | Atherosclerosis of nonautologous biological coronary artery bypass graft(s) with other forms of angina pectoris |
|  | I25.739 | Atherosclerosis of nonautologous biological coronary artery bypass graft(s) with unspecified angina pectoris |
|  | I25.750 | Atherosclerosis of native coronary artery of transplanted heart with unstable angina |
|  | I25.760 | Atherosclerosis of bypass graft of coronary artery of transplanted heart with unstable angina |
|  | I25.790 | Atherosclerosis of other coronary artery bypass graft(s) with unstable angina pectoris |
|  | I25.791 | Atherosclerosis of other coronary artery bypass graft(s) with angina pectoris with documented spasm |
|  | I25.798 | Atherosclerosis of other coronary artery bypass graft(s) with other forms of angina pectoris |
|  | I25.799 | Atherosclerosis of other coronary artery bypass graft(s) with unspecified angina pectoris |
|  | I25.8 | Other forms of chronic ischemic heart disease |
|  | I25.810 | Atherosclerosis of coronary artery bypass graft(s) without angina pectoris |
|  | I25.82 | Chronic total occlusion of coronary artery |
|  | I25.83 | Coronary atherosclerosis due to lipid rich plaque |
|  | I25.84 | Coronary atherosclerosis due to calcified coronary lesion |
|  | I25.89 | Other forms of chronic ischemic heart disease |
|  | I25.9 | Chronic ischemic heart disease, unspecified |
|  | Z95.1 | Presence of aortocoronary bypass graft |
|  | Z95.5 | Presence of coronary angioplasty implant and graft |
|  | Z98.6 | Angioplasty status |
|  | Z98.61 | Coronary angioplasty status |
| **ICD-9 CM Procedure Codes** | 36.01 | Single vessel percutaneous transluminal coronary angioplasty [PTCA] or coronary atherectomy without mention of thrombolytic agent |
|  | 36.02 | Single vessel percutaneous transluminal coronary angioplasty [PTCA] or coronary atherectomy with mention of thrombolytic agent |
|  | 36.03 | Open Chest Coronary Artery Angioplasty |
|  | 36.05 | Multiple vessel percutaneous transluminal coronary angioplasty [PTCA] or coronary atherectomy performed during the same operation, with or without mention of thrombolytic agent |
|  | 36.06 | Insertion of non-drug-eluting coronary artery stent(s) |
|  | 36.07 | Insertion of drug-eluting coronary artery stent(s) |
|  | 36.09 | Other Removal Of Coronary Artery Obstruction |
|  | 36.1 | Aortocoronary Bypass For Heart Revascularization, Not Otherwise Specified |
|  | 36.11 | (Aorto)coronary bypass of one coronary artery |
|  | 36.12 | Aortocoronary Bypass Of Two Coronary Arteries |
|  | 36.13 | Aortocoronary Bypass Of Three Coronary Arteries |
|  | 36.14 | Aortocoronary Bypass Of Four Or More Coronary Arteries |
|  | 36.15 | Single Internal Mammary-Coronary Artery Bypass |
|  | 36.16 | Double Internal Mammary-Coronary Artery Bypass |
|  | 36.17 | Abdominal-Coronary Artery Bypass |
|  | 36.19 | Other Bypass Anastomosis For Heart Revascularization |
|  | 36.2 | CABG surgery |
| **ICD-10 Procedure Codes** | 021008C | Bypass Coronary Artery, One Artery from Thoracic Artery with Zooplastic Tissue, Open Approach |
|  | 021008W | Bypass Coronary Artery, One Artery from Aorta with Zooplastic Tissue, Open Approach |
|  | 210088 | Bypass Coronary Artery, One Artery from Right Internal Mammary with Zooplastic Tissue, Open Approach |
|  | 210089 | Bypass Coronary Artery, One Artery from Left Internal Mammary with Zooplastic Tissue, Open Approach |
|  | 210093 | Bypass Coronary Artery, One Artery from Coronary Artery with Autologous Venous Tissue, Open Approach |
|  | 210098 | Bypass Coronary Artery, One Artery from Right Internal Mammary with Autologous Venous Tissue, Open Approach |
|  | 210099 | Bypass Coronary Artery, One Artery from Left Internal Mammary with Autologous Venous Tissue, Open Approach |
|  | 021009C | Bypass Coronary Artery, One Artery from Thoracic Artery with Autologous Venous Tissue, Open Approach |
|  | 021009F | Bypass Coronary Artery, One Artery from Abdominal Artery with Autologous Venous Tissue, Open Approach |
|  | 021009W | Bypass Coronary Artery, One Artery from Aorta with Autologous Venous Tissue, Open Approach |
|  | 02100A3 | Bypass Coronary Artery, One Artery from Coronary Artery with Autologous Arterial Tissue, Open Approach |
|  | 02100A8 | Bypass Coronary Artery, One Artery from Right Internal Mammary with Autologous Arterial Tissue, Open Approach |
|  | 02100A9 | Bypass Coronary Artery, One Artery from Left Internal Mammary with Autologous Arterial Tissue, Open Approach |
|  | 02100AC | Bypass Coronary Artery, One Artery from Thoracic Artery with Autologous Arterial Tissue, Open Approach |
|  | 02100AF | Bypass Coronary Artery, One Artery from Abdominal Artery with Autologous Arterial Tissue, Open Approach |
|  | 02100AW | Bypass Coronary Artery, One Artery from Aorta with Autologous Arterial Tissue, Open Approach |
|  | 02100J3 | Bypass Coronary Artery, One Artery from Coronary Artery with Synthetic Substitute, Open Approach |
|  | 02100J8 | Bypass Coronary Artery, One Artery from Right Internal Mammary with Synthetic Substitute, Open Approach |
|  | 02100J9 | Bypass Coronary Artery, One Artery from Left Internal Mammary with Synthetic Substitute, Open Approach |
|  | 02100JC | Bypass Coronary Artery, One Artery from Thoracic Artery with Synthetic Substitute, Open Approach |
|  | 02100JF | Bypass Coronary Artery, One Artery from Abdominal Artery with Synthetic Substitute, Open Approach |
|  | 02100JW | Bypass Coronary Artery, One Artery from Aorta with Synthetic Substitute, Open Approach |
|  | 02100K3 | Bypass Coronary Artery, One Artery from Coronary Artery with Nonautologous Tissue Substitute, Open Approach |
|  | 02100K8 | Bypass Coronary Artery, One Artery from Right Internal Mammary with Nonautologous Tissue Substitute, Open Approach |
|  | 02100K9 | Bypass Coronary Artery, One Artery from Left Internal Mammary with Nonautologous Tissue Substitute, Open Approach |
|  | 02100KC | Bypass Coronary Artery, One Artery from Thoracic Artery with Nonautologous Tissue Substitute, Open Approach |
|  | 02100KF | Bypass Coronary Artery, One Artery from Abdominal Artery with Nonautologous Tissue Substitute, Open Approach |
|  | 02100KW | Bypass Coronary Artery, One Artery from Aorta with Nonautologous Tissue Substitute, Open Approach |
|  | 02100Z3 | Bypass Coronary Artery, One Artery from Coronary Artery, Open Approach |
|  | 02100Z8 | Bypass Coronary Artery, One Artery from Right Internal Mammary, Open Approach |
|  | 02100Z9 | Bypass Coronary Artery, One Artery from Left Internal Mammary, Open Approach |
|  | 02100ZC | Bypass Coronary Artery, One Artery from Thoracic Artery, Open Approach |
|  | 02100ZF | Bypass Coronary Artery, One Artery from Abdominal Artery, Open Approach |
|  | 210488 | Bypass Coronary Artery, One Artery from Right Internal Mammary with Zooplastic Tissue, Percutaneous Endoscopic Approach |
|  | 021048C | Bypass Coronary Artery, One Artery from Thoracic Artery with Zooplastic Tissue, Percutaneous Endoscopic Approach |
|  | 021048W | Bypass Coronary Artery, One Artery from Aorta with Zooplastic Tissue, Percutaneous Endoscopic Approach |
|  | 210489 | Bypass Coronary Artery, One Artery from Left Internal Mammary with Zooplastic Tissue, Percutaneous Endoscopic Approach |
|  | 210493 | Bypass Coronary Artery, One Artery from Coronary Artery with Autologous Venous Tissue, Percutaneous Endoscopic Approach |
|  | 210498 | Bypass Coronary Artery, One Artery from Right Internal Mammary with Autologous Venous Tissue, Percutaneous Endoscopic Approach |
|  | 210499 | Bypass Coronary Artery, One Artery from Left Internal Mammary with Autologous Venous Tissue, Percutaneous Endoscopic Approach |
|  | 021049C | Bypass Coronary Artery, One Artery from Thoracic Artery with Autologous Venous Tissue, Percutaneous Endoscopic Approach |
|  | 021049F | Bypass Coronary Artery, One Artery from Abdominal Artery with Autologous Venous Tissue, Percutaneous Endoscopic Approach |
|  | 021049W | Bypass Coronary Artery, One Artery from Aorta with Autologous Venous Tissue, Percutaneous Endoscopic Approach |
|  | 02104A3 | Bypass Coronary Artery, One Artery from Coronary Artery with Autologous Arterial Tissue, Percutaneous Endoscopic Approach |
|  | 02104A8 | Bypass Coronary Artery, One Artery from Right Internal Mammary with Autologous Arterial Tissue, Percutaneous Endoscopic Approach |
|  | 02104A9 | Bypass Coronary Artery, One Artery from Left Internal Mammary with Autologous Arterial Tissue, Percutaneous Endoscopic Approach |
|  | 02104AC | Bypass Coronary Artery, One Artery from Thoracic Artery with Autologous Arterial Tissue, Percutaneous Endoscopic Approach |
|  | 02104AF | Bypass Coronary Artery, One Artery from Abdominal Artery with Autologous Arterial Tissue, Percutaneous Endoscopic Approach |
|  | 02104AW | Bypass Coronary Artery, One Artery from Aorta with Autologous Arterial Tissue, Percutaneous Endoscopic Approach |
|  | 02104J3 | Bypass Coronary Artery, One Artery from Coronary Artery with Synthetic Substitute, Percutaneous Endoscopic Approach |
|  | 02104J8 | Bypass Coronary Artery, One Artery from Right Internal Mammary with Synthetic Substitute, Percutaneous Endoscopic Approach |
|  | 02104J9 | Bypass Coronary Artery, One Artery from Left Internal Mammary with Synthetic Substitute, Percutaneous Endoscopic Approach |
|  | 02104JC | Bypass Coronary Artery, One Artery from Thoracic Artery with Synthetic Substitute, Percutaneous Endoscopic Approach |
|  | 02104JF | Bypass Coronary Artery, One Artery from Abdominal Artery with Synthetic Substitute, Percutaneous Endoscopic Approach |
|  | 02104JW | Bypass Coronary Artery, One Artery from Aorta with Synthetic Substitute, Percutaneous Endoscopic Approach |
|  | 02104K3 | Bypass Coronary Artery, One Artery from Coronary Artery with Nonautologous Tissue Substitute, Percutaneous Endoscopic Approach |
|  | 02104K8 | Bypass Coronary Artery, One Artery from Right Internal Mammary with Nonautologous Tissue Substitute, Percutaneous Endoscopic Approach |
|  | 02104K9 | Bypass Coronary Artery, One Artery from Left Internal Mammary with Nonautologous Tissue Substitute, Percutaneous Endoscopic Approach |
|  | 02104KC | Bypass Coronary Artery, One Artery from Thoracic Artery with Nonautologous Tissue Substitute, Percutaneous Endoscopic Approach |
|  | 02104KF | Bypass Coronary Artery, One Artery from Abdominal Artery with Nonautologous Tissue Substitute, Percutaneous Endoscopic Approach |
|  | 02104KW | Bypass Coronary Artery, One Artery from Aorta with Nonautologous Tissue Substitute, Percutaneous Endoscopic Approach |
|  | 02104Z3 | Bypass Coronary Artery, One Artery from Coronary Artery, Percutaneous Endoscopic Approach |
|  | 02104Z8 | Bypass Coronary Artery, One Artery from Right Internal Mammary, Percutaneous Endoscopic Approach |
|  | 02104Z9 | Bypass Coronary Artery, One Artery from Left Internal Mammary, Percutaneous Endoscopic Approach |
|  | 02104ZC | Bypass Coronary Artery, One Artery from Thoracic Artery, Percutaneous Endoscopic Approach |
|  | 02104ZF | Bypass Coronary Artery, One Artery from Abdominal Artery, Percutaneous Endoscopic Approach |
|  | 021108W | Bypass Coronary Artery, Two Arteries from Aorta with Zooplastic Tissue, Open Approach |
|  | 211098 | Bypass Coronary Artery, Two Arteries from Right Internal Mammary with Autologous Venous Tissue, Open Approach |
|  | 211099 | Bypass Coronary Artery, Two Arteries from Left Internal Mammary with Autologous Venous Tissue, Open Approach |
|  | 021109C | Bypass Coronary Artery, Two Arteries from Thoracic Artery with Autologous Venous Tissue, Open Approach |
|  | 021109W | Bypass Coronary Artery, Two Arteries from Aorta with Autologous Venous Tissue, Open Approach |
|  | 02110A8 | Bypass Coronary Artery, Two Arteries from Right Internal Mammary with Autologous Arterial Tissue, Open Approach |
|  | 02110A9 | Bypass Coronary Artery, Two Arteries from Left Internal Mammary with Autologous Arterial Tissue, Open Approach |
|  | 02110AC | Bypass Coronary Artery, Two Arteries from Thoracic Artery with Autologous Arterial Tissue, Open Approach |
|  | 02110AW | Bypass Coronary Artery, Two Arteries from Aorta with Autologous Arterial Tissue, Open Approach |
|  | 02110J8 | Bypass Coronary Artery, Two Arteries from Right Internal Mammary with Synthetic Substitute, Open Approach |
|  | 02110J9 | Bypass Coronary Artery, Two Arteries from Left Internal Mammary with Synthetic Substitute, Open Approach |
|  | 02110JC | Bypass Coronary Artery, Two Arteries from Thoracic Artery with Synthetic Substitute, Open Approach |
|  | 02110JW | Bypass Coronary Artery, Two Arteries from Aorta with Synthetic Substitute, Open Approach |
|  | 02110K8 | Bypass Coronary Artery, Two Arteries from Right Internal Mammary with Nonautologous Tissue Substitute, Open Approach |
|  | 02110K9 | Bypass Coronary Artery, Two Arteries from Left Internal Mammary with Nonautologous Tissue Substitute, Open Approach |
|  | 02110KC | Bypass Coronary Artery, Two Arteries from Thoracic Artery with Nonautologous Tissue Substitute, Open Approach |
|  | 02110KW | Bypass Coronary Artery, Two Arteries from Aorta with Nonautologous Tissue Substitute, Open Approach |
|  | 02110Z8 | Bypass Coronary Artery, Two Arteries from Right Internal Mammary, Open Approach |
|  | 02110Z9 | Bypass Coronary Artery, Two Arteries from Left Internal Mammary, Open Approach |
|  | 02110ZC | Bypass Coronary Artery, Two Arteries from Thoracic Artery, Open Approach |
|  | 021148W | Bypass Coronary Artery, Two Arteries from Aorta with Zooplastic Tissue, Percutaneous Endoscopic Approach |
|  | 211498 | Bypass Coronary Artery, Two Arteries from Right Internal Mammary with Autologous Venous Tissue, Percutaneous Endoscopic Approach |
|  | 211499 | Bypass Coronary Artery, Two Arteries from Left Internal Mammary with Autologous Venous Tissue, Percutaneous Endoscopic Approach |
|  | 021149C | Bypass Coronary Artery, Two Arteries from Thoracic Artery with Autologous Venous Tissue, Percutaneous Endoscopic Approach |
|  | 021149W | Bypass Coronary Artery, Two Arteries from Aorta with Autologous Venous Tissue, Percutaneous Endoscopic Approach |
|  | 02114A8 | Bypass Coronary Artery, Two Arteries from Right Internal Mammary with Autologous Arterial Tissue, Percutaneous Endoscopic Approach |
|  | 02114A9 | Bypass Coronary Artery, Two Arteries from Left Internal Mammary with Autologous Arterial Tissue, Percutaneous Endoscopic Approach |
|  | 02114AC | Bypass Coronary Artery, Two Arteries from Thoracic Artery with Autologous Arterial Tissue, Percutaneous Endoscopic Approach |
|  | 02114AW | Bypass Coronary Artery, Two Arteries from Aorta with Autologous Arterial Tissue, Percutaneous Endoscopic Approach |
|  | 02114J8 | Bypass Coronary Artery, Two Arteries from Right Internal Mammary with Synthetic Substitute, Percutaneous Endoscopic Approach |
|  | 02114J9 | Bypass Coronary Artery, Two Arteries from Left Internal Mammary with Synthetic Substitute, Percutaneous Endoscopic Approach |
|  | 02114JC | Bypass Coronary Artery, Two Arteries from Thoracic Artery with Synthetic Substitute, Percutaneous Endoscopic Approach |
|  | 02114JW | Bypass Coronary Artery, Two Arteries from Aorta with Synthetic Substitute, Percutaneous Endoscopic Approach |
|  | 02114K8 | Bypass Coronary Artery, Two Arteries from Right Internal Mammary with Nonautologous Tissue Substitute, Percutaneous Endoscopic Approach |
|  | 02114K9 | Bypass Coronary Artery, Two Arteries from Left Internal Mammary with Nonautologous Tissue Substitute, Percutaneous Endoscopic Approach |
|  | 02114KC | Bypass Coronary Artery, Two Arteries from Thoracic Artery with Nonautologous Tissue Substitute, Percutaneous Endoscopic Approach |
|  | 02114KW | Bypass Coronary Artery, Two Arteries from Aorta with Nonautologous Tissue Substitute, Percutaneous Endoscopic Approach |
|  | 02114Z8 | Bypass Coronary Artery, Two Arteries from Right Internal Mammary, Percutaneous Endoscopic Approach |
|  | 02114Z9 | Bypass Coronary Artery, Two Arteries from Left Internal Mammary, Percutaneous Endoscopic Approach |
|  | 02114ZC | Bypass Coronary Artery, Two Arteries from Thoracic Artery, Percutaneous Endoscopic Approach |
|  | 021209C | Bypass Coronary Artery, Three Arteries from Thoracic Artery with Autologous Venous Tissue, Open Approach |
|  | 021209W | Bypass Coronary Artery, Three Arteries from Aorta with Autologous Venous Tissue, Open Approach |
|  | 02120AC | Bypass Coronary Artery, Three Arteries from Thoracic Artery with Autologous Arterial Tissue, Open Approach |
|  | 02120AW | Bypass Coronary Artery, Three Arteries from Aorta with Autologous Arterial Tissue, Open Approach |
|  | 02120JC | Bypass Coronary Artery, Three Arteries from Thoracic Artery with Synthetic Substitute, Open Approach |
|  | 02120JW | Bypass Coronary Artery, Three Arteries from Aorta with Synthetic Substitute, Open Approach |
|  | 02120KC | Bypass Coronary Artery, Three Arteries from Thoracic Artery with Nonautologous Tissue Substitute, Open Approach |
|  | 02120KW | Bypass Coronary Artery, Three Arteries from Aorta with Nonautologous Tissue Substitute, Open Approach |
|  | 02120ZC | Bypass Coronary Artery, Three Arteries from Thoracic Artery, Open Approach |
|  | 021249C | Bypass Coronary Artery, Three Arteries from Thoracic Artery with Autologous Venous Tissue, Percutaneous Endoscopic Approach |
|  | 021249W | Bypass Coronary Artery, Three Arteries from Aorta with Autologous Venous Tissue, Percutaneous Endoscopic Approach |
|  | 02124AC | Bypass Coronary Artery, Three Arteries from Thoracic Artery with Autologous Arterial Tissue, Percutaneous Endoscopic Approach |
|  | 02124AW | Bypass Coronary Artery, Three Arteries from Aorta with Autologous Arterial Tissue, Percutaneous Endoscopic Approach |
|  | 02124JC | Bypass Coronary Artery, Three Arteries from Thoracic Artery with Synthetic Substitute, Percutaneous Endoscopic Approach |
|  | 02124JW | Bypass Coronary Artery, Three Arteries from Aorta with Synthetic Substitute, Percutaneous Endoscopic Approach |
|  | 02124KC | Bypass Coronary Artery, Three Arteries from Thoracic Artery with Nonautologous Tissue Substitute, Percutaneous Endoscopic Approach |
|  | 02124KW | Bypass Coronary Artery, Three Arteries from Aorta with Nonautologous Tissue Substitute, Percutaneous Endoscopic Approach |
|  | 02124ZC | Bypass Coronary Artery, Three Arteries from Thoracic Artery, Percutaneous Endoscopic Approach |
|  | 021308W | Bypass Coronary Artery, Four or More Arteries from Aorta with Zooplastic Tissue, Open Approach |
|  | 021309C | Bypass Coronary Artery, Four or More Arteries from Thoracic Artery with Autologous Venous Tissue, Open Approach |
|  | 021309W | Bypass Coronary Artery, Four or More Arteries from Aorta with Autologous Venous Tissue, Open Approach |
|  | 02130AC | Bypass Coronary Artery, Four or More Arteries from Thoracic Artery with Autologous Arterial Tissue, Open Approach |
|  | 02130AW | Bypass Coronary Artery, Four or More Arteries from Aorta with Autologous Arterial Tissue, Open Approach |
|  | 02130JC | Bypass Coronary Artery, Four or More Arteries from Thoracic Artery with Synthetic Substitute, Open Approach |
|  | 02130JW | Bypass Coronary Artery, Four or More Arteries from Aorta with Synthetic Substitute, Open Approach |
|  | 02130KC | Bypass Coronary Artery, Four or More Arteries from Thoracic Artery with Nonautologous Tissue Substitute, Open Approach |
|  | 02130KW | Bypass Coronary Artery, Four or More Arteries from Aorta with Nonautologous Tissue Substitute, Open Approach |
|  | 02130ZC | Bypass Coronary Artery, Four or More Arteries from Thoracic Artery, Open Approach |
|  | 021349C | Bypass Coronary Artery, Four or More Arteries from Thoracic Artery with Autologous Venous Tissue, Percutaneous Endoscopic Approach |
|  | 021349W | Bypass Coronary Artery, Four or More Arteries from Aorta with Autologous Venous Tissue, Percutaneous Endoscopic Approach |
|  | 02134AC | Bypass Coronary Artery, Four or More Arteries from Thoracic Artery with Autologous Arterial Tissue, Percutaneous Endoscopic Approach |
|  | 02134AW | Bypass Coronary Artery, Four or More Arteries from Aorta with Autologous Arterial Tissue, Percutaneous Endoscopic Approach |
|  | 02134JC | Bypass Coronary Artery, Four or More Arteries from Thoracic Artery with Synthetic Substitute, Percutaneous Endoscopic Approach |
|  | 02134JW | Bypass Coronary Artery, Four or More Arteries from Aorta with Synthetic Substitute, Percutaneous Endoscopic Approach |
|  | 02134KC | Bypass Coronary Artery, Four or More Arteries from Thoracic Artery with Nonautologous Tissue Substitute, Percutaneous Endoscopic Approach |
|  | 02134KW | Bypass Coronary Artery, Four or More Arteries from Aorta with Nonautologous Tissue Substitute, Percutaneous Endoscopic Approach |
|  | 02134ZC | Bypass Coronary Artery, Four or More Arteries from Thoracic Artery, Percutaneous Endoscopic Approach |
|  | 0270xxx | Dilation of Coronary Artery, One Artery |
|  | 02700ZZ | Dilation of Coronary Artery, One Artery, Open Approach |
|  | 02703ZZ | Dilation of Coronary Artery, One Artery, Percutaneous Approach. |
|  | 02704ZZ | Dilation of Coronary Artery, One Artery, Percutaneous Endoscopic Approach |
|  | 0271xxx | Dilation of Coronary Artery, Two Arteries |
|  | 02710ZZ | Dilation of Coronary Artery, Two Arteries, Open Approach |
|  | 02713ZZ | Dilation of Coronary Artery, Two Arteries, Percutaneous Approach |
|  | 02714ZZ | Dilation of Coronary Artery, Two Arteries, Percutaneous Endoscopic Approach |
|  | 0272xxx | Dilation of Coronary Artery, Three Arteries |
|  | 02720ZZ | Dilation of Coronary Artery, Three Arteries, Open Approach |
|  | 02723ZZ | Dilation of Coronary Artery, Three Arteries, Percutaneous Approach |
|  | 02724ZZ | Dilation of Coronary Artery, Three Arteries, Percutaneous Endoscopic Approach |
|  | 0273xxx | Dilation of Coronary Artery, Four or More Arteries |
|  | 02730ZZ | Dilation of Coronary Artery, Four or More Arteries, Open Approach |
|  | 02733ZZ | Dilation of Coronary Artery, Four or More Arteries, Percutaneous Approach |
|  | 02734ZZ | Dilation of Coronary Artery, Four or More Arteries, Percutaneous Endoscopic Approach |
|  | 02C0xxx | Extirpation of Matter from Coronary Artery |
|  | 02C00Z6 | Extirpation of Matter from Coronary Artery, One Artery, Bifurcation, Open Approach |
|  | 02C00ZZ | Extirpation of Matter from Coronary Artery, One Artery, Open Approach |
|  | 02C03Z6 | Extirpation of Matter from Coronary Artery, One Artery, Bifurcation, Percutaneous Approach |
|  | 02C03Z7 | Extirpation of Matter from Coronary Artery, One Artery, Orbital Atherectomy Technique, Percutaneous Approach |
|  | 02C03ZZ | Extirpation of Matter from Coronary Artery, One Artery, Percutaneous Approach |
|  | 02C04Z6 | Extirpation of Matter from Coronary Artery, One Artery, Bifurcation, Percutaneous Endoscopic Approach |
|  | 02C04ZZ | Extirpation of Matter from Coronary Artery, One Artery, Percutaneous Endoscopic Approach |
|  | 02C1xxx | Extirpation of Matter from Coronary Artery, Two Arteries |
|  | 02C10Z6 | Extirpation of Matter from Coronary Artery, Two Artery, Bifurcation, Open Approach |
|  | 02C10ZZ | Extirpation of Matter from Coronary Artery, Two Arteries, Open Approach |
|  | 02C13Z6 | Extirpation of Matter from Coronary Artery, Two Arteries, Bifurcation, Percutaneous Approach |
|  | 02C13Z7 | Extirpation of Matter from Coronary Artery, Two Arteries, Orbital Atherectomy Technique, Percutaneous Approach |
|  | 02C13ZZ | Extirpation of Matter from Coronary Artery, Two Arteries, Percutaneous Approach |
|  | 02C14Z6 | Extirpation of Matter from Coronary Artery, Two Artery, Bifurcation, Percutaneous Endoscopic Approach |
|  | 02C14ZZ | Extirpation of Matter from Coronary Artery, Two Arteries, Percutaneous Endoscopic Approach |
|  | 02C2xxx | Extirpation of Matter from Coronary Artery, Three Arteries |
|  | 02C20Z6 | Extirpation of Matter from Coronary Artery, Three Arteries, Bifurcation, Open Approach |
|  | 02C20ZZ | Extirpation of Matter from Coronary Artery, Three Arteries, Open Approach |
|  | 02C23Z6 | Extirpation of Matter from Coronary Artery, Three Artery, Bifurcation, Open Approach |
|  | 02C23Z7 | Extirpation of Matter from Coronary Artery, Three Arteries, Open Approach |
|  | 02C23ZZ | Extirpation of Matter from Coronary Artery, Three Arteries, Percutaneous Approach |
|  | 02C24Z6 | Extirpation of Matter from Coronary Artery, Three Arteries, Bifurcation, Percutaneous Endoscopic Approach |
|  | 02C24ZZ | Extirpation of Matter from Coronary Artery, Three Arteries, Percutaneous Endoscopic Approach |
|  | 02C3xxx | Extirpation of Matter from Coronary Artery, Four or More Arteries |
|  | 02C30Z6 | Extirpation of Matter from Coronary Artery, Four or More Arteries, Bifurcation, Open Approach |
|  | 02C30ZZ | Extirpation of Matter from Coronary Artery, Four or More Arteries, Open Approach |
|  | 02C33Z6 | Extirpation of Matter from Coronary Artery, Four or More Arteries, Bifurcation, Open Approach |
|  | 02C33Z7 | Extirpation of Matter from Coronary Artery, Four or More Arteries, Percutaneous Endoscopic Approach |
|  | 02C33ZZ | Extirpation of Matter from Coronary Artery, Four or More Arteries, Percutaneous Approach |
|  | 02C34Z6 | Extirpation of Matter from Coronary Artery, Four or More Arteries, Bifurcation, Percutaneous Endoscopic Approach |
|  | 02C34ZZ | Extirpation of Matter from Coronary Artery, Four or More Arteries, Percutaneous Endoscopic Approach |
|  | 3E07017 | Introduction of Other Thrombolytic into Coronary Artery, Open Approach |
|  | 3E070PZ | Introduction of Platelet Inhibitor into Coronary Artery, Open Approach |
|  | 3E07317 | Introduction of Other Thrombolytic into Coronary Artery, Percutaneous Approach |
|  | 3E073PZ | Introduction of Platelet Inhibitor into Coronary Artery, Percutaneous Approach |
| **CPT**  **Codes** | 33510 | Coronary artery bypass, vein only; single coronary venous graft |
|  | 33511 | Coronary artery bypass, vein only; 2 coronary venous grafts |
|  | 33512 | Coronary artery bypass, vein only; 3 coronary venous grafts |
|  | 33513 | Coronary artery bypass, vein only; 4 coronary venous grafts |
|  | 33514 | Coronary artery bypass, vein only; 5 coronary venous grafts |
|  | 33516 | Coronary artery bypass, vein only; 6 or more coronary venous grafts |
|  | 33517 | Coronary artery bypass, using venous graft(s) and arterial graft(s); single vein graft |
|  | 33518 | Coronary artery bypass, using venous graft(s) and arterial graft(s); 2 venous grafts |
|  | 33519 | Coronary artery bypass, using venous graft(s) and arterial graft(s); 3 venous grafts |
|  | 33521 | Coronary artery bypass, using venous graft(s) and arterial graft(s); 4 venous grafts |
|  | 33522 | Coronary artery bypass, using venous graft(s) and arterial graft(s); 5 venous grafts |
|  | 33523 | Coronary artery bypass, using venous graft(s) and arterial graft(s); 6 or more venous grafts |
|  | 33533 | Coronary artery bypass, using arterial graft(s); single arterial graft |
|  | 33534 | Coronary artery bypass, using arterial graft(s); 2 coronary arterial grafts |
|  | 33535 | Coronary artery bypass, using arterial graft(s); 3 coronary arterial grafts |
|  | 33536 | Coronary artery bypass, using arterial graft(s); 4 or more coronary arterial grafts |
|  | 33545 | Arterial Grafting for Coronary Artery Bypass |
|  | 33572 | Coronary Endarterectomy Procedures |
|  | 35500 | Vein Bypass Graft Procedures |
|  | 92920 | Percutaneous revascularization services performed for occlusive disease of the coronary vessels |
|  | 92921 | Percutaneous transluminal coronary angioplasty; each additional branch of a major coronary artery |
|  | 92924 | Percutaneous transluminal coronary atherectomy, with coronary angioplasty when performed; single major coronary artery or branch |
|  | 92925 | Percutaneous transluminal coronary atherectomy, with coronary angioplasty when performed; each additional branch of a major coronary artery |
|  | 92928 | Percutaneous transcatheter placement of intracoronary stent(s), with coronary angioplasty when performed; single major coronary artery or branch |
|  | 92929 | Percutaneous transcatheter placement of intracoronary stent(s), with coronary angioplasty when performed; each additional branch of a major coronary artery |
|  | 92933 | Percutaneous transluminal coronary atherectomy, with intracoronary stent, with coronary angioplasty when performed; single major coronary artery or branch |
|  | 92934 | Percutaneous transluminal coronary atherectomy, with intracoronary stent, with coronary angioplasty when performed; each additional branch of a major coronary artery |
|  | 92937 | Percutaneous transluminal revascularization of or through coronary artery bypass graft (internal mammary, free arterial, venous), any combination of intracoronary stent, atherectomy and angioplasty, including distal protection when performed; single vessel |
|  | 92938 | Percutaneous transluminal revascularization of or through coronary artery bypass graft (internal mammary, free arterial, venous), any combination of intracoronary stent, atherectomy and angioplasty, including distal protection when performed; each additional branch subtended by the bypass graft |
|  | 92941 | Percutaneous transluminal revascularization of acute total/subtotal occlusion during acute myocardial infarction, coronary artery or coronary artery bypass graft, any combination of intracoronary stent, atherectomy and angioplasty, including aspiration thrombectomy when performed, single vessel |
|  | 92943 | Percutaneous transluminal revascularization of acute total/subtotal occlusion during acute myocardial infarction, coronary artery or coronary artery bypass graft, any combination of intracoronary stent, atherectomy and angioplasty, including aspiration thrombectomy when performed, single vessel |
|  | 92944 | Percutaneous transluminal revascularization of chronic total occlusion, coronary artery, coronary artery branch, or coronary artery bypass graft, any combination of intracoronary stent, atherectomy and angioplasty; each additional coronary artery, coronary artery branch, or bypass graft |
|  | 92973 | Percutaneous transluminal coronary thrombectomy mechanical |
|  | 92974 | Transcatheter placement of radiation delivery device for subsequent coronary intravascular brachytherapy |
|  | 92975 | Thrombolysis, coronary; by intracoronary infusion, including selective coronary angiography |
|  | 92977 | An intravenous injection or infusion of a thrombolytic agent |
|  | 92980 | Transcatheter placement of an intracoronary stent(s), percutaneous, with or without other therapeutic intervention, any method; single vessel |
|  | 92981 | Transcatheter placement of an intracoronary stent(s), percutaneous, with or without other therapeutic intervention, any method; each additional vessel |
|  | 92982 | Percutaneous transluminal coronary balloon angioplasty; single vessel |
|  | 92984 | Percutaneous transluminal coronary balloon angioplasty; each additional vessel |
|  | 92995 | Percutaneous transluminal coronary atherectomy, by mechanical or other method, with or without balloon angioplasty; single vessel |
|  | 92996 | Percutaneous transluminal coronary atherectomy, by mechanical or other method, with or without balloon angioplasty; each additional vessel |
|  | 92998 | Percutaneous transluminal pulmonary artery balloon angioplasty. |
|  | C9600 | Percutaneous transcatheter placement of drug eluting intracoronary stent(s), with coronary angioplasty when performed; single major coronary artery or branch |
|  | C9601 | Percutaneous transcatheter placement of drug-eluting intracoronary stent(s), with coronary angioplasty when performed; each additional branch of a major coronary artery |
|  | C9602 | Percutaneous transluminal coronary atherectomy, with drug eluting intracoronary stent, with coronary angioplasty when performed; single major coronary artery or branch |
|  | C9603 | Percutaneous transluminal coronary atherectomy, with drug-eluting intracoronary stent, with coronary angioplasty when performed; each additional branch of a major coronary artery |
|  | C9604 | Percutaneous transluminal revascularization of or through coronary artery bypass graft (internal mammary, free arterial, venous), any combination of drug-eluting intracoronary stent, atherectomy and angioplasty, including distal protection when performed; single vessel |
|  | C9605 | Percutaneous transluminal revascularization of or through coronary artery bypass graft (internal mammary, free arterial, venous), any combination of drug-eluting intracoronary stent, atherectomy and angioplasty, including distal protection when performed; each additional branch subtended by the bypass graft |
|  | C9606 | Percutaneous transluminal revascularization of acute total/subtotal occlusion during acute myocardial infarction, coronary artery or coronary artery bypass graft, any combination of drug-eluting intracoronary stent, atherectomy and angioplasty, including aspiration thrombectomy when performed, single vessel |
|  | C9607 | Percutaneous transluminal revascularization of chronic total occlusion, coronary artery, coronary artery branch, or coronary artery bypass graft, any combination of drug-eluting intracoronary stent, atherectomy and angioplasty; single vessel |
|  | C9608 | Percutaneous transluminal revascularization of chronic total occlusion, coronary artery, coronary artery branch, or coronary artery bypass graft, any combination of drug-eluting intracoronary stent, atherectomy and angioplasty; each additional coronary artery, coronary artery branch, or bypass graft |
| **SNOMED Condition Codes** | 282006 | Acute myocardial infarction of other lateral wall episode of care unspecified |
|  | 1077002 | Acute myocardial infarction of other specified sites episode of care unspecified |
|  | 1755008 | Old myocardial infarction |
|  | 3098007 | Acute myocardial infarction of other specified sites episode of care unspecified |
|  | 10273003 | Acute myocardial infarction of other specified sites episode of care unspecified |
|  | 15990001 | Acute myocardial infarction of other lateral wall episode of care unspecified |
|  | 22298006 | Heart attack |
|  | 52035003 | Acute myocardial infarction of other anterior wall episode of care unspecified |
|  | 54329005 | Anterior myocardial infarction NOS |
|  | 56276002 | Acute myocardial infarction of other specified sites episode of care unspecified |
|  | 57054005 | Acute myocardial infarction |
|  | 58612006 | Acute myocardial infarction of lateral wall |
|  | 59063002 | Acute myocardial infarction of other lateral wall episode of care unspecified |
|  | 62695002 | Acute myocardial infarction of other anterior wall episode of care unspecified |
|  | 64627002 | Acute myocardial infarction of other lateral wall episode of care unspecified |
|  | 65547006 | Acute inferolateral infarction |
|  | 70211005 | Acute anterolateral infarction |
|  | 70422006 | Acute subendocardial infarction |
|  | 73795002 | Inferior myocardial infarction NOS |
|  | 76593002 | Acute myocardial infarction of inferoposterior wall |
|  | 79009004 | Acute myocardial infarction of other specified sites episode of care unspecified |
|  | 164868007 | Acute myocardial infarction of other anterior wall episode of care unspecified |
|  | 164871004 | Acute myocardial infarction of other lateral wall episode of care unspecified |
|  | 194809007 | Acute myocardial infarction of other specified sites episode of care unspecified |
|  | 194856005 | Acute myocardial infarction of unspecified site episode of care unspecified |
|  | 194857001 | Acute myocardial infarction of unspecified site episode of care unspecified |
|  | 194858006 | Acute myocardial infarction of unspecified site episode of care unspecified |
|  | 194862000 | Acute myocardial infarction of unspecified site episode of care unspecified |
|  | 194866002 | Acute myocardial infarction of unspecified site episode of care unspecified |
|  | 194868001 | Acute myocardial infarction of unspecified site episode of care unspecified |
|  | 233825009 | Acute myocardial infarction of other anterior wall episode of care unspecified |
|  | 233826005 | Acute myocardial infarction of other anterior wall episode of care unspecified |
|  | 233827001 | Acute Q wave infarction - anterolateral |
|  | 233829003 | Acute myocardial infarction of other inferior wall episode of care unspecified |
|  | 233830008 | Acute myocardial infarction of other inferior wall episode of care unspecified |
|  | 233831007 | Acute Q wave infarction - inferolateral |
|  | 233833005 | Acute myocardial infarction of other lateral wall episode of care unspecified |
|  | 233834004 | Acute myocardial infarction of other lateral wall episode of care unspecified |
|  | 233838001 | Posterior myocardial infarction NOS |
|  | 304914007 | Acute Q-wave infarct |
|  | 311792005 | Acute myocardial infarction of other anterior wall episode of care unspecified |
|  | 311793000 | Acute myocardial infarction of other inferior wall episode of care unspecified |
|  | 371817007 | Acute myocardial infarction of other specified sites episode of care unspecified |
|  | 394710008 | Acute myocardial infarction of unspecified site episode of care unspecified |
|  | 401303003 | Acute ST segment elevation myocardial infarction |
|  | 401314000 | Acute non-ST segment elevation myocardial infarction |
|  | 418044006 | Acute myocardial infarction of unspecified site episode of care unspecified |
|  | 703164000 | Acute myocardial infarction of other anterior wall episode of care unspecified |
|  | 703165004 | Acute myocardial infarction of other anterior wall episode of care unspecified |
|  | 703209002 | Acute myocardial infarction of other inferior wall episode of care unspecified |
|  | 703210007 | Acute myocardial infarction of other anterior wall episode of care unspecified |
|  | 703211006 | Acute myocardial infarction of unspecified site episode of care unspecified |
|  | 703213009 | Acute myocardial infarction of other inferior wall episode of care unspecified |
|  | 703251009 | Acute myocardial infarction of inferior wall involving right ventricle |
|  | 703252002 | Acute myocardial infarction of other anterior wall episode of care unspecified |
|  | 703253007 | Acute myocardial infarction of other inferior wall episode of care unspecified |
|  | 840309000 | Acute ST segment elevation myocardial infarction due to proximal left anterior descending coronary artery occlusion |
|  | 840312002 | Acute ST segment elevation myocardial infarction due to mid left anterior descending coronary artery occlusion |
|  | 840316004 | Acute ST segment elevation myocardial infarction due to distal left anterior descending coronary artery occlusion |
|  | 840609007 | Acute ST segment elevation myocardial infarction due to occlusion of anterior descending branch of left coronary artery |
|  | 840680009 | Acute ST segment elevation myocardial infarction due to occlusion of septal branch of anterior descending branch of left coronary artery |
|  | 846668006 | Acute ST segment elevation myocardial infarction due to occlusion of diagonal branch of anterior descending branch of left coronary artery |
|  | 846683001 | Acute ST segment elevation myocardial infarction due to occlusion of intermediate artery |
|  | 868214006 | Acute ST segment elevation myocardial infarction due to occlusion of proximal portion of right coronary artery |
|  | 868217004 | Acute ST segment elevation myocardial infarction due to occlusion of distal portion of right coronary artery |
|  | 868220007 | Acute ST segment elevation myocardial infarction due to occlusion of mid portion of right coronary artery |
|  | 868224003 | Acute ST segment elevation myocardial infarction due to occlusion of marginal branch of right coronary artery |
|  | 868225002 | Acute ST segment elevation myocardial infarction due to occlusion of posterior descending branch of right coronary artery |
|  | 868226001 | Acute ST segment elevation myocardial infarction due to occlusion of posterior lateral branch of right coronary artery |
|  | 896689003 | Acute myocardial infarction due to occlusion of circumflex branch of left coronary artery |
|  | 896691006 | Acute ST segment elevation myocardial infarction due to occlusion of circumflex branch of left coronary artery |
|  | 23311000119105 | Acute myocardial infarction due to right coronary artery occlusion |
|  | 12238111000119106 | Acute ST segment elevation myocardial infarction of inferoposterior wall |
|  | 15713081000119108 | Acute ST segment elevation myocardial infarction due to left coronary artery occlusion |
|  | 15962541000119106 | Acute ST segment elevation myocardial infarction of anteroapical wall |
|  | 703164000 | Acute anterior ST segment elevation myocardial infarction |
|  | 62695002 | Acute anteroseptal myocardial infarction |
|  | 723860000 | Arrhythmia due to and following acute myocardial infarction |
|  | 194863005 | Atrial septal defect due to and following acute myocardial infarction |
|  | 233847009 | Cardiac rupture due to and following acute myocardial infarction |
|  | 827164008 | Delayed postmyocardial infarction pericarditis |
|  | 827163002 | Early postmyocardial infarction pericarditis |
|  | 194862000 | Hemopericardium due to and following acute myocardial infarction |
|  | 1142308000 | Mitral valve regurgitation due to acute myocardial infarction |
|  | 703330009 | Mitral valve regurgitation due to acute myocardial infarction with papillary muscle and chordal rupture |
|  | 42531007 | Other acute and subacute forms of ischaemic heart disease |
|  | 46109009 | Other acute and subacute forms of ischaemic heart disease |
|  | 194823009 | Other acute and subacute forms of ischaemic heart disease |
|  | 194849004 | Other acute and subacute forms of ischaemic heart disease |
|  | 413439005 | Other acute and subacute forms of ischaemic heart disease |
|  | 713405002 | Other acute and subacute forms of ischaemic heart disease |
|  | 71023004 | Pericarditis secondary to acute myocardial infarction |
|  | 314116003 | Post infarct angina |
|  | 233929004 | Post-infarction mural thrombus |
|  | 233885007 | Post-infarction pericarditis |
|  | 233846000 | Post-infarction ventricular septal defect |
|  | 66189004 | Postmyocardial infarction syndrome |
|  | 70422006 | Subendocardial infarction episode of care unspecified |
|  | 703360004 | Subendocardial infarction episode of care unspecified |
|  | 194856005 | Subsequent myocardial infarction |
|  | 194858006 | Subsequent myocardial infarction of inferior wall |
|  | 703360004 | Subsequent non-ST segment elevation myocardial infarction |
|  | 703211006 | Subsequent ST segment elevation myocardial infarction |
|  | 703209002 | Subsequent ST segment elevation myocardial infarction of inferior wall |
|  | 16415081000119104 | Supraventricular tachycardia following acute myocardial infarction |
|  | 194868001 | Thrombosis of atrium, auricular appendage, and ventricle due to and following acute myocardial infarction |
|  | 233838001 | True posterior wall infarction episode of care unspecified |
|  | 723858002 | Ventricular aneurysm due to and following acute myocardial infarction |
|  | 233889001 | Post-infarction hemopericardium |
|  | 129574000 | Postoperative myocardial infarction |
|  | 311792005 | Postoperative transmural myocardial infarction of anterior wall |
|  | 311793000 | Postoperative transmural myocardial infarction of inferior wall |
|  | 233889001 | Post-infarction hemopericardium |
|  | 429245005 | Recurrent coronary arteriosclerosis after percutaneous transluminal coronary angioplasty |
|  | 194865003 | Rupture of cardiac wall without hemopericardium as current complication following acute myocardial infarction |
|  | 194866002 | Rupture of chordae tendinae due to and following acute myocardial infarction |
|  | 16528621000119102 | Rupture of interventricular septum following acute myocardial infarction |
|  | 30277009 | Rupture of ventricle due to acute myocardial infarction |
|  | 386138005 | Stented coronary artery |
| **SNOMED Procedure Codes** | 3546002 | Coronary artery venous graft |
|  | 11101003 | Coronary Angioplasty |
|  | 14201006 | Open angioplasty of coronary artery |
|  | 41339005 | Coronary angioplasty |
|  | 68466008 | Percut transluminal balloon angioplasty one coronary artery |
|  | 85053006 | Percutaneous transluminal coronary angioplasty, multiple vessels |
|  | 175021005 | Allograft bypass of coronary artery |
|  | 175029007 | Prosthetic replacement of coronary artery |
|  | 232717009 | Coronary artery bypass graft |
|  | 232720001 | Coronary artery bypass grafts x 2 |
|  | 232721002 | Coronary artery bypass grafts x 3 |
|  | 232722009 | Coronary artery bypass grafts x 4 |
|  | 399261000 | [V]Presence of coronary artery bypass graft |
|  | 429639007 | Perc translum balloon angioplasty insert 1-2 stents cor art |
|  | 707828002 | Percut translum cutting balloon angioplasty coronary artery |
|  | 736968006 | Saphenous vein graft replacement of three coronary arteries |
|  | 736969003 | Saphenous vein graft replacement of two coronary arteries |
|  | 736973000 | Allograft replacement of two coronary arteries |
|  | 61236006 | Aortocoronary artery bypass graft repeated |
|  | 737276005 | Aortocoronary bypass graft present |
|  | 131521000119101 | Bare metal stent in anterior descending branch of left coronary artery |
|  | 131561000119106 | Bare metal stent in branch of right coronary artery |
|  | 131591000119104 | Bare metal stent in circumflex branch of left coronary artery |
|  | 286241000119106 | Bare metal stent in posterior descending branch of right coronary artery |
|  | 286261000119105 | Bypass stent graft present |
|  | 286271000119104 | Coronary artery bypass graft stent present |
|  | 737278006 | Coronary artery graft present |
|  | 251019006 | Coronary bypass graft finding |
|  | 131601000119106 | Drug coated stent in anterior descending branch of left coronary artery |
|  | 131551000119109 | Drug coated stent in branch of right coronary artery |
|  | 131581000119102 | Drug coated stent in circumflex branch of left coronary artery |
|  | 286251000119108 | Drug coated stent in posterior descending branch of right coronary artery |
|  | 473155009 | History of angioplasty |
|  | 429117009 | History of aortofemoral bypass surgery |
|  | 429367008 | History of arterial bypass of lower limb artery |
|  | 399261000 | History of coronary artery bypass grafting |
|  | 428308007 | History of percutaneous transluminal coronary angioplasty |
|  | 428375006 | History of placement of stent for coronary artery disease |
|  | 428912006 | History of placement of stent in anterior descending branch of left coronary artery |
|  | 428664003 | History of placement of stent in circumflex branch of left coronary artery |
|  | 130541000119100 | History of placement of stent in coronary artery bypass graft |
|  | 371822007 | Patient post percutaneous transluminal coronary angioplasty |
|  | 216621000119100 | Stent in anterior descending branch of left coronary artery |
|  | 130551000119103 | Stent in branch of right coronary artery |
|  | 216631000119102 | Stent in circumflex branch of left coronary artery |
|  | 286281000119101 | Stent in posterior descending branch of right coronary artery |
