## Supplementary material for "Impact of Participation Bias on Disease Prevalence Estimation in the *All of Us* Research Program: A Case Study of Ischemic Heart Disease and Stroke": eTable 2

#### **eTable 2.** Effect estimates from a multivariate logistic regression model predicting completion of the Personal Medical History (PMH) survey among participants with linked EHR.

|  | **OR (95% CI)** |
| --- | --- |
| **Age at enrollment** (per 1 SD increase) | 1.09 (1.08-1.11) |
| **Sex assigned at** birth [Reference: Female] |  |
| Male at birth | 0.68 (0.67-0.70) |
| Not male, not female, refused, skipped, or no matching concept | 1.20 (1.11-1.29) |
| **Born in non-US countries** [Reference: Born in the US] | 0.93 (0.90-0.96) |
| **Self-reported race** [Reference: Non-Hispanic White] |  |
| Hispanic or Latino | 0.50 (0.48-0.51) |
| NH Asian | 0.60 (0.57-0.64) |
| NH Black or African American | 0.37 (0.36-0.38) |
| NH More than one population | 0.81 (0.76-0.86) |
| **Marital status** [Reference: Married/Living with partner] |  |
| Divorced/Separated/Widowed | 0.85 (0.83-0.87) |
| Never married | 0.99 (0.96-1.01) |
| **Has no health insurance** [Reference: Has health insurance] | 0.87 (0.83-0.90) |
| **Current homeownership** [Reference: Own] |  |
| Other arrangement | 0.61 (0.59-0.64) |
| Rent | 0.69 (0.67-0.70) |
| **Less than college education** [Reference: College or higher] | 0.54 (0.52-0.55) |
| **Not currently employed for wages** [Reference: Currently employed] | 0.93 (0.91-0.95) |
| **Household income < 50k** [Reference: 50k or higher] | 0.63 (0.62-0.65) |
| **Lifetime alcohol consumption** [Reference: Never] | 1.45 (1.41-1.50) |
| **Lifetime cigarette smoking** [Reference: Never] | 0.79 (0.77-0.80) |
| **Not at all/A little bit/Somewhat confident filling out medical forms** [Reference: Extremely/Quite a bit] | 0.64 (0.62-0.66) |
| **Often/Always having problems learning about your medical condition** [Reference: Never/Occasionally/Sometime] | 0.80 (0.77-0.83) |
| **Often/Always needing help reading health-related materials** [Reference: Never/Occasionally/Sometime] | 0.91 (0.87-0.96) |

#### Abbreviation: EHR, electronic health records; OR, odds ratio; CI, confidence interval.
