## Supplementary material for "Impact of Participation Bias on Disease Prevalence Estimation in the *All of Us* Research Program: A Case Study of Ischemic Heart Disease and Stroke": eTable 3

#### **e****Table 3.** Prevalence estimates for ischemic heart disease (IHD) and stroke by case definition and weighting scheme among 124,192 PMH survey respondents.

| **Phenotype** | **Sample** | **Case definition** | **Weighting scheme** | **Prevalence estimate (95% CI)** |
| --- | --- | --- | --- | --- |
| Ischemic  heart  disease | Overall | EHR-based | - | 6.4 (6.3-6.5) |
|  | Non-respondents | EHR-based | - | 7.2 (7.0-7.3) |
|  | Respondents | EHR-based | - | 5.6 (5.4-5.7) |
|  |  | PMH-based | Unweighted | 5.9 (5.7-6.0) |
|  |  |  | Poststratification* | 6.0 (5.8-6.1) |
| Stroke | Overall | EHR-based | - | 2.8 (2.7-2.8) |
|  | Non-respondents | EHR-based | - | 3.3 (3.2-3.4) |
|  | Respondents | EHR-based | - | 2.2 (2.1-2.3) |
|  |  | PMH-based | Unweighted | 3.6 (3.5-3.7) |
|  |  |  | Poststratification* | 3.8 (3.6-3.9) |

* We applied poststratification weights estimated using annual household income and educational attainment.

Abbreviation: EHR, electronic health records; CI, confidence interval.
