## Supplementary material for "Impact of Participation Bias on Disease Prevalence Estimation in the *All of Us* Research Program: A Case Study of Ischemic Heart Disease and Stroke": eTable 4

**eTable 4.** Prevalence estimates for ischemic heart disease and stroke by case definition, weighting scheme, and self-reported race and ethnicity.

| **Phenotype** | **Weighting scheme** | **Self-reported race and ethnicity** | **Prevalence estimate (95% CI)** |
| --- | --- | --- | --- |
| Ischemic  heart  disease (IHD) | Unweighted | NH Asian | 2.8 (2.3-3.4) |
|  |  | NH Black or African American | 4.5 (4.1-4.8) |
|  |  | Hispanic or Latino | 3.0 (2.7-3.3) |
|  |  | NH More than one population | 3.6 (2.9-4.4) |
|  |  | Refused/Skipped/None of these | 6.9 (6.2-7.6) |
|  |  | NH White | 6.7 (6.5-6.8) |
|  | Poststratification | NH Asian | 2.8 (2.2-3.3) |
|  |  | NH Black or African American | 4.8 (4.3-5.2) |
|  |  | Hispanic or Latino | 3.2 (2.9-3.5) |
|  |  | NH More than one population | 3.6 (2.7-4.5) |
|  |  | Refused/Skipped/None of these | 6.7 (6.0-7.3) |
|  |  | NH White | 7.0 (6.8-7.2) |
| Stroke | Unweighted | NH Asian | 1.5 (1.1-1.9) |
|  |  | NH Black or African American | 4.7 (4.3-5.1) |
|  |  | Hispanic or Latino | 2.0 (1.8-2.2) |
|  |  | NH More than one population | 2.9 (2.2-3.6) |
|  |  | Refused/Skipped/None of these | 3.8 (3.3-4.3) |
|  |  | NH White | 3.7 (3.6-3.9) |
|  | Poststratification | NH Asian | 1.5 (1.0-1.9) |
|  |  | NH Black or African American | 4.7 (4.3-5.1) |
|  |  | Hispanic or Latino | 2.0 (1.7-2.2) |
|  |  | NH More than one population | 3.1 (2.3-4.0) |
|  |  | Refused/Skipped/None of these | 3.8 (3.2-4.3) |
|  |  | NH White | 4.1 (3.9-4.2) |

Abbreviation: NH, non-Hispanic; CI, confidence interval.
